# Effectiveness of COVID-19 mRNA primary and booster vaccination against infection and hospitalisation during pregnancy: a target trial emulation and meta-analysis of data from 4 European countries

**DOI:** 10.1101/2025.05.16.25327593

**Authors:** Núria Mercade-Besora, Berta Raventós, Nhung TH Trinh, Huiqi Li, Saeed Hayati, Agustina Giuliodori, Theresa Burkard, Edward Burn, Martí Català, Daniel Prieto-Alhambra, Daniel R. Morales, Fredrik Nyberg, Hedvig Marie Egeland Nordeng, Talita Duarte-Salles

**Author notes:** joint first authors (Núria Mercade-Besora, Berta Raventós). joint senior authors (Hedvig Marie Egeland Nordeng, Talita Duarte-Salles). deceased. **Corresponding author:** Talita Duarte-Salles Fundació Institut Universitari per a la recerca a l’Atenció Primària de Salut Jordi Gol i Gurina (IDIAPJGol), Gran Via Corts Catalanes, 587 àtic, 08007 Barcelona – Spain.

## Abstract

**Objective:** To estimate vaccine effectiveness (VE) of COVID-19 mRNA vaccines administered during pregnancy. Primary (2-dose) vaccination was compared with unvaccinated women; and 1^st^ booster (3rd dose) with primary (2-dose) vaccination.

**Design:** Target trial emulation cohort study.

**Setting:** Routinely collected health data from UK, Sweden, Spain, and Norway. All had complete vaccine exposure data and were mapped to the OMOP common data model.

**Participants:** Pregnant women (aged 12-55) with ≤34 weeks of gestation, no history of COVID-19 in the 90 days prior and ≥1 year of data visibility. Women were matched 1:1 to those with no dose and 2-dose vaccination, respectively, using weekly sequential matching from vaccination roll-out to 2023 (Swedish data: 2024).

**Main outcome measures:** The main outcomes were COVID-19 infection, hospitalisation and Intensive Care Unit (ICU) admission (COVID-related, excluding delivery-related care). Hazard ratios (HR) were estimated with Cox regression from 15 days post-vaccination, and VE was reported as 100 x (1-HR). Analyses were stratified by pregnancy trimester and vaccine brand. Database-specific results were pooled using a random-effects meta-analysis.

**Results:** We included 62,061 and 41,096 pregnant women at date of vaccination with a first and third dose, respectively. Median follow-up across databases ranged from 42 to 55 days for primary vaccination and 73 to 116 days for boosters. For primary vaccination, the meta-analysis showed a VE of 26.2% (16.2 to 35.1; I^2^ =0.7) against infection and 55.6% (44.1 to 64.8; I^2^ =0) against hospitalisation. For boosters, corresponding figures were 33.6% (-1.6 to 56.6; I^2^=1) and 34.1% (0.6 to 56.4; I^2^ =0.5). Event counts were insufficient to analyse VE against ICU admission. Results were consistent across pregnancy trimesters and vaccine brands.

**Conclusions:** Vaccination with a primary (2-dose) or a 1^st^ booster (3rd dose) during pregnancy was effective in preventing COVID-19 hospitalisation, with modest and heterogeneous VE against infection across 4 European countries.

**Summary:** *What is already known on this topic:*

- Pregnant women are at increased risk of severe complications from COVID-19, which can affect both mothers and their infants.
- Vaccination during pregnancy has been shown to be safe and effective, with benefits of vaccination outweighing any potential risks.
- Studies examining the effectiveness of COVID-19 vaccines during pregnancy have primarily focused on primary vaccination with BNT162b2, with limited follow-up, sample size and representativeness.

*What this study adds:*

- This study provides evidence on the effectiveness of COVID-19 mRNA vaccines administered during pregnancy using routinely collected data of over 124,122 pregnancies from 4 European countries.
- Primary (2-dose) vaccination against COVID-19 during pregnancy was effective in preventing infection during pregnancy with a vaccine effectiveness of 26%, and 55% for preventing COVID-19 related hospitalisations.
- A 1^st^ booster (3^rd^ dose) had a vaccine effectiveness of 30-35% against COVID-19 infection and COVID-19-related hospitalisation compared to primary (2-dose) vaccination.

## INTRODUCTION

Pregnancy is associated with an increased risk of severe complications from coronavirus disease 2019 (COVID-19). Pregnant women with COVID-19 are at increased risk of admission to intensive care units (ICU) and receiving invasive ventilation, compared to non-pregnant women with COVID-19.(1) They are also more likely to experience severe pregnancy complications, including preeclampsia, preterm birth and stillbirth compared to pregnant women without COVID-19.(1, 2) Given the risks faced by both pregnant patients and neonates, pregnant women were included in vaccination campaigns before clinical trials on this population were completed. The lack of pregnancy-specific evidence when the COVID-19 vaccination campaigns started led to diminished vaccine confidence, leaving many pregnant women unvaccinated,(3, 4) and at an increased risk of severe outcomes. Unclear messaging and vaccine hesitancy has likely contributed to perpetuating risks for unvaccinated pregnant women, with COVID-19 persisting among the leading causes of maternal deaths in the United Kingdom (UK) since the pandemic started.(5–7)

Current evidence suggests that COVID-19 vaccines administered during pregnancy are safe (8–13) and have similar effectiveness to that observed in the general population.(11, 14, 15) However, existing observational studies on the effectiveness of COVID-19 vaccines during pregnancy have focused on BNT162b2 (Pfizer-BioNTech),(16, 17) with only one study examining the effectiveness of a third dose against severe COVID-19 disease,(17) and none assessing the effectiveness of mRNA-1273 (Moderna) in real-world settings. In addition, pregnancy-specific evidence has largely been derived from single-database studies with short follow-up and limited representativeness. Further data are still needed to complement these findings and to assess vaccine effectiveness (VE) during pregnancy in the evolving pandemic context. Data on booster doses is particularly relevant given that many women have already been vaccinated before entering pregnancy, and given current recommendations endorsed by the World Health Organization that now advise a vaccine dose during pregnancy, regardless of prior vaccination.(18)

The objective of this study was to estimate the clinical effectiveness of BNT162b2 and mRNA-1273 vaccines used for primary (2-dose) and for 1^st^ booster (3^rd^ dose) vaccination during pregnancy in preventing COVID-19 infection and COVID-19-related admission to hospitals or ICUs.

## METHODS

### Design: target trial specifications

We specified two target trials to evaluate the effectiveness of BNT162b2 and mRNA-1273 vaccines administered during pregnancy in preventing COVID-19 infection, hospitalisation, and ICU admissions (Supplementary Table 1). The first target trial aimed to evaluate the effectiveness of primary (2-dose) vaccination, with second doses administered within 17 to 42 days and 24 to 49 days after a first dose of BNT162b2 and mRNA-1273, respectively. Dosage intervals allowed in the study between first and second doses were based on reasonable variation around the 6 weeks following recommendations of clinical studies when first approved.(19, 20) The second target trial aimed to evaluate the effectiveness of the first booster vaccination, defined by the administration of a third dose given during pregnancy among women who had received the second dose at least three months prior, regardless of whether this occurred during or before pregnancy.

### Data sources

The specified target trials were emulated using four routine healthcare databases with complete vaccination coverage from Norway, Spain, Sweden, and the UK. Data from Norway and Sweden were obtained from nation-wide registries: the Norwegian Linked Health Registries (NLHR) at the University of Oslo (data from Norway is referred to as UiO from now onwards), and the Swedish Covid-19 Investigation for Future Insights – a Population Epidemiology Approach using Register Linkage (SCIFI-PEARL) project. Data from Spain and the UK were obtained from primary care electronic health records (EHR). Spanish data was obtained from the Information System for Research in Primary Care (SIDIAP) database, which captures more than 75% of the population living in Catalonia, Spain, and has linkage to hospital discharge data.(21, 22) Data from the UK was obtained from the Clinical Practice Research Datalink (CPRD) GOLD (23), which covers approximately 19% of the UK population across all UK nations. A detailed description of these RWD sources is provided in Supplementary Text 1. Notably, all data sources had complete records of vaccination. Pregnancy data was provided directly by the data source, through linkage, or inferred. For UiO and SCIFI-PEARL, pregnancy data was obtained through linkage to the Medical Birth Registry of Norway and the Swedish Birth Registry, which had information on pregnancies lasting longer than 12 and 22 weeks, respectively. We used the UiO pregnancy algorithm to identify pregnancies that ended before gestational week 12 and to complement those notified to the Medical Birth Registry of Norway.(24) No linkage to birth registries was employed for SIDIAP, which provided detailed pregnancy information from reproductive health clinics, or for CPRD GOLD, where pregnancy episodes were inferred from primary care data using a validated algorithm.(25) All databases were mapped to the Observational Medical Outcomes Partnership (OMOP) Common Data Model (CDM), therefore allowing federated analyses without sharing of person-level data. Pregnancy episodes were mapped to the perinatal expansion table in the OMOP CDM, which facilitates the identification of pregnancy episodes in OMOP-formatted data.(26)

### Study population

We defined the study population according to target trial eligibility criteria. Both trials were designed to include pregnant women aged 12 to 55 years with gestational age less than 34 weeks. We excluded participants who, on inclusion, had tested positive or had a clinical diagnosis of COVID-19 in their records in the 90 days prior, and those who did not have data visibility of at least 365 days before pregnancy start.

### Outcomes

Outcomes were COVID-19 infection, COVID-19-related hospitalisation, and admission to the ICU. COVID-19 infection cases were identified based on positive severe acute respiratory syndrome coronavirus 2 (SARS-CoV-2) test results or diagnosis with a compatible clinical code. COVID-19-related hospitalisation was defined as an admission with a compatible clinical code or positive SARS-CoV-2 test within 21 days before to 3 days after admission. The same criteria were used for identifying ICU admission with COVID-19, as described in prior studies.(21) We restricted hospitalisations to those unrelated to delivery by excluding hospitalisations occurring within 3 days of delivery (i.e. ± 3 days of delivery date). Information on hospitalisations was not available in CPRD GOLD, and ICU admissions were only available in SCIFI-PEARL (up to 20/11/2022) and SIDIAP.

### Follow up and covariate assessment

We conducted a sequential matching based on calendar week to derive exposed and unexposed cohorts in each data source. Each week, pregnant women receiving the vaccine dose of interest on that week were matched to unexposed pregnant women. Missing vaccination records were interpreted as the pregnant women not having received a vaccine. On each trial week, the variables used to apply the inclusion and exclusion criteria, the matching variables, and baseline covariates were measured at week start day. For the first trial, vaccinated pregnant women (from now on called the exposed arm) were matched in a 1:1 ratio with pregnant women who were not vaccinated that week (from now on called the unexposed arm). Exact matching was based on age at pregnancy start date (2-year band) and gestational age (4-week band), defined as the date of the first day in the last menstrual period. Matching was also based on propensity scores with a 0.2 caliper width. Maternal information for calculating propensity scores included: demographics (age, geographical region, days of prior observation), health-related factors (underlying conditions and number of prior pregnancies), and health-seeking behavior (number of health visits, number of prior SARS-CoV-2 tests, and history of prior influenza and tetanus, diphtheria and pertussis vaccinations). The same strategy was applied for the second trial, with the exposed arm being pregnant women receiving a third dose of an mRNA COVID-19 vaccine and the unexposed arm being matched pregnant women with completed primary (2-dose) vaccination schema. Here, the time since the second dose of the primary schema (in 1-month bands) was also used for exact matching. Detailed information on the matching variables and time windows used for its assessment can be found in the Supplementary Figure 1. The matching was repeated on each subsequent week, from the start of the vaccination roll-out in each country (December 2020) until 90 days before the end of the study period, defined as the end of data availability of each database, which was end of 2023 for all databases except for SCIFI-PEARL, which had data up to end of 2024 (see Supplementary Text 1).

Time zero (index date) for each matched pair was defined as the date of vaccination. This was the date of the first dose for the first trial, and the date of the third dose for the second trial. We defined the end of follow-up as the earliest of the following: the end of pregnancy, the date when the individual was lost to follow up or died, and the occurrence of an outcome of interest. Follow-up for each matched pair was censored if one of pair experienced any of the reasons above except for outcome occurrence. Follow-up for each matched pair was also censored if any of its members were not adherent to the treatment strategy assigned at baseline. This included: a change in the vaccination status of the unexposed counterpart (e.g. receiving a first dose for the analysis of VE of primary vaccination, receiving a third dose for the analysis of booster VE), receiving a subsequent booster dose for the exposed counterpart (e.g. receiving a third dose for the analysis of VE of primary vaccination), or not receiving a second dose in the pre-specified time window for the analysis of VE of primary vaccination. Women initially assigned to the unexposed group who received a vaccine during follow-up were allowed to re-enter the study in the exposed arm from that time point and be matched to a new unexposed counterpart.

### Statistical analysis

Absolute standardised mean differences (SMD) were used to evaluate observed confounding, with an absolute standardised mean difference ≤ 0.1 interpreted as balanced/comparable across arms. We then used 70 negative control outcomes (NCO) to assess the presence of residual (unobserved) confounding. Of those, 43 NCOs were pre-specified based on previous methodological research,(27) and 27 were incorporated after review by a clinical epidemiologist. We used the Kaplan-Meier estimator to construct cumulative incidence curves and to estimate the risk for each outcome. Risk of study outcomes were compared using hazard ratios (HR) derived from Cox proportional hazard models.(28) Given that unexposed women could be re-enrolled as exposed if vaccinated, the Huber (sandwich) estimator was applied to obtain 95% confidence intervals (CI). We analysed outcomes in the full population and in subgroups defined by strata of vaccine brand and pregnancy trimester. VE (%) was estimated as 100x (1 – HR) and was assessed from 15 days after vaccination until end of follow-up. Additionally, VE was also assessed in different follow-up periods: 0 to 14, 15 to 28, 29 to 90, 91 to 180 and 181 to 365 days.

As a secondary analysis, we did not consider the end of pregnancy as a timepoint to conclude follow-up. We conducted several sensitivity analyses to test the robustness of our findings. For COVID-19 infection, we restricted COVID-19 cases to those with a record of a positive SARS-CoV-2 test result. For COVID-19 hospitalisation, we removed the exclusion of delivery-related care and included hospitalisations occurring within 3 days of the delivery date.

Random effects meta-analyses across databases were conducted to obtain overall effect estimates when estimates were available from at least three databases. Heterogeneity between database-specific results was assessed using the I^2^ statistic. Event counts with ≤ 5 occurrences were obscured to preserve privacy. All analyses were conducted using R (version 4.2.3). All analytical code, code lists for variables (outcomes, COVID-19 tests, diagnoses, etc.) and NCOs are publicly available in GitHub (https://github.com/rwepi-idiapjgol/COVIDVaccineEffectivenessDuringPregnancy).

### Patient and Public Involvement

No patients or members of the public were directly involved in the design or analysis of the reported data.

## RESULTS

### Study population

A total of 305,415 pregnant women met the eligibility criteria for the first trial (i.e. primary [2-dose] vaccination compared to unvaccinated) and 198,269 for the analysis of the second trial (i.e. 1^st^ booster (3^rd^ dose) compared to primary [2-dose] vaccination). For the first trial, 62,061 vaccinated pregnant women (CPRD GOLD: 2,741; SCIFI-PEARL: 36,398; SIDAP: 7,479; UiO: 15,443) were matched 1:1 with the same number of unexposed pregnant women (Figure 1, Supplementary Figure 2-4). For the second trial, 41,096 pregnant women vaccinated with a third dose (CPRD GOLD: 1,892; SCIFI-PEARL: 26,195; SIDAP: 2,218; UiO: 10,791) were matched 1:1 to pregnant women eligible to receive a third dose at the time of matching (Figure 1, Supplementary Fig 2-4). The proportion of enrolled controls later re-enrolled as exposed ranged from 7 to 18% across analyses (Supplementary Table 2).

**Figure 1.**
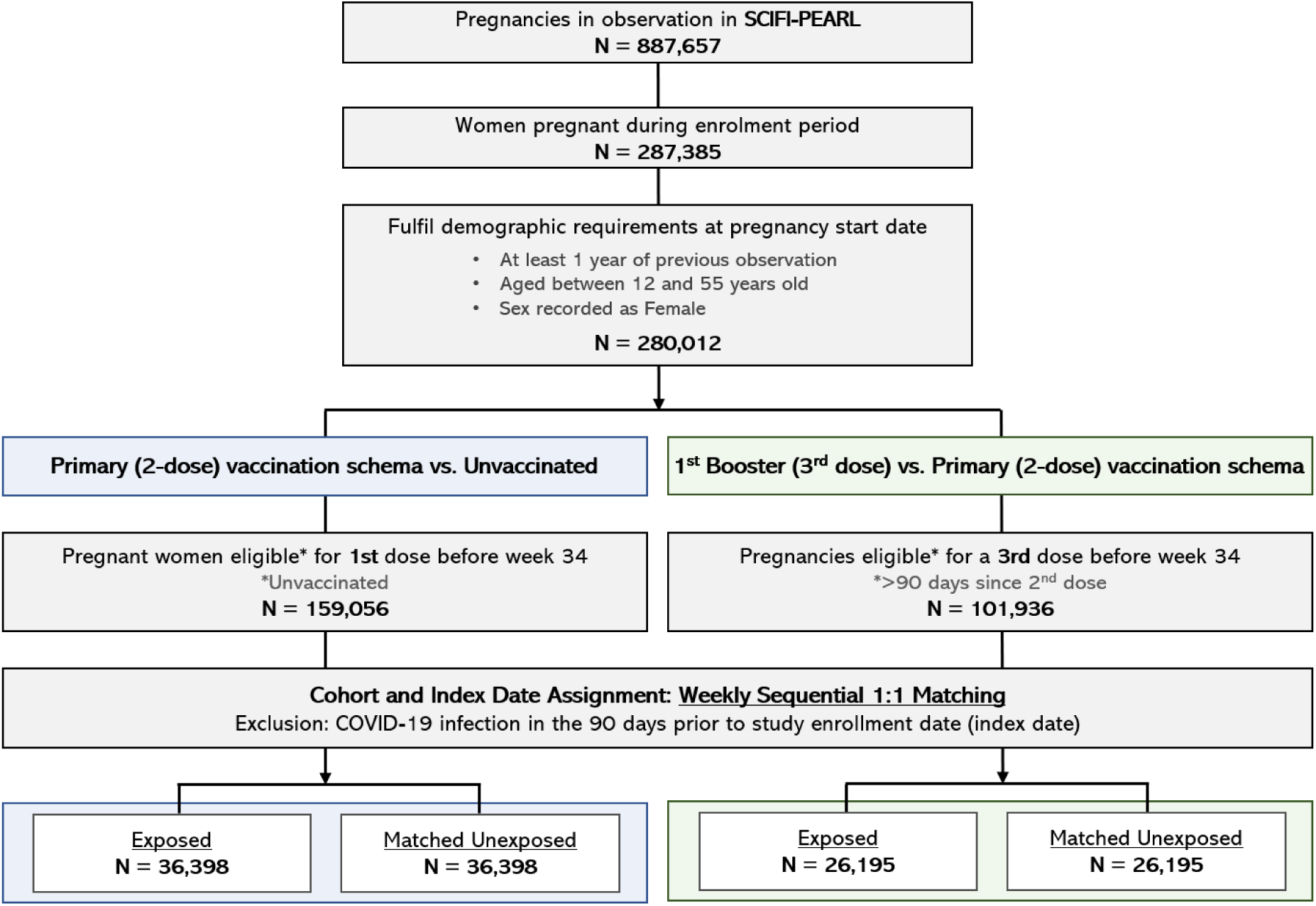
Study population and flow chart for SCIFI-PEARL.

Baseline characteristics were similar between the exposed and unexposed groups for both trials (Table 1-2). The median age of participants was approximately 31-34 years old. Most women were vaccinated during the first and second trimester, while 11-26% of women were vaccinated during the third trimester. Over 80% of women were vaccinated with a first dose of BNT162b2 across all databases except for UiO (66%). BNT162b2 was administered for 80% of women receiving a 1^st^ booster (3^rd^ dose) except for CPRD GOLD (64.7%) and SIDIAP (30.3%). Extended tables characterising the study populations at baseline, with additional medical conditions and prior medication use, can be found in Supplementary Tables 3-10. SMDs were suggestive of covariate balance, and can be explored in the interactive web App with all results (https://dpa-pde-oxford.shinyapps.io/COVID19VaccineEffectivenessDuringPregnancy/).

**Table 1.** Baseline characteristics of matched pregnant women included for the analysis of VE of primary (2-dose) vaccination vs. unvaccinated.

|  | **CPRD GOLD** |  | **SCIFI-PEARL** |  | **SIDIAP** |  | **Uio** |  |
| --- | --- | --- | --- | --- | --- | --- | --- | --- |
| Variable name | Exposed | Matched unexposed | Exposed | Matched unexposed | Exposed | Matched unexposed | Exposed | Matched unexposed |
| Number of pregnancies, N | 2,741 | 2,741 | 36,398 | 36,398 | 7,479 | 7,479 | 15,443 | 15,443 |
| Age, Median [Q25 - Q75] | 32 [28 - 35] | 32 [28 - 35] | 31 [28 - 35] | 31 [28 - 35] | 32 [28 - 36] | 32 [28 - 36] | 31 [28 - 34] | 31 [28 - 34] |
| First-dose vaccine brand, N (%) |  |  |  |  |  |  |  |  |
| BNT162b2 | 2,461 (89.8%) | - | 29,050 (79.8%) | - | 5,983 (80.0%) | - | 10,195 (66.0%) | - |
| mRNA-1273 | 280 (10.2%) | - | 7,348 (20.2%) | - | 1,496 (20.0%) | - | 5,248 (34.0%) | - |
| Trimester at index date, N (%) |  |  |  |  |  |  |  |  |
| Trimester 1 | 1,107 (40.4%) | 1,107 (40.4%) | 9,441 (25.9%) | 9,441 (25.9%) | 2,183 (29.2%) | 2,183 (29.2%) | 5,201 (33.7%) | 5,201 (33.7%) |
| Trimester 2 | 954 (34.8%) | 954 (34.8%) | 18,782 (51.6%) | 18,782 (51.6%) | 3,351 (44.8%) | 3,351 (44.8%) | 6,421 (41.6%) | 6,421 (41.6%) |
| Trimester 3 | 680 (24.8%) | 680 (24.8%) | 8,175 (22.5%) | 8,175 (22.5%) | 1,945 (26.0%) | 1,945 (26.0%) | 3,821 (24.7%) | 3,821 (24.7%) |
| Health visits*, Median [Q25 - Q75] | 24 [14 - 42] | 20 [11 - 38] | 8 [2- 22] | 4 [1 - 14] | 32 [16- 52] | 20 [10- 36] | 30 [16 - 48] | 20 [11- 34] |
| Conditions**, N (%) |  |  |  |  |  |  |  |  |
| Asthma | 212 (7.7%) | 173 (6.3%) | 1,778 (4.9%) | 1,502 (4.1%) | 455 (6.1%) | 370 (4.9%) | 2,207 (14.3%) | 2,004 (13.0%) |
| Chronic hypertension | 30 (1.1%) | 28 (1.0%) | 1,267 (3.5%) | 995 (2.7%) | 178 (2.4%) | 155 (2.1%) | 1,122 (7.3%) | 1,049 (6.8%) |
| COPD | 0 (0.0%) | 0 (0.0%) | 13 (0.0%) | 6 (0.0%) | 8 (0.1%) | 6 (0.1%) | 75 (0.5%) | 74 (0.5%) |
| Diabetes | 110 (4.0%) | 94 (3.4%) | 1,017 (2.8%) | 983 (2.7%) | 445 (5.9%) | 405 (5.4%) | 823 (5.3%) | 826 (5.3%) |
| Obesity | 287 (10.5%) | 254 (9.3%) | 2,691 (7.4%) | 2,671 (7.3%) | 1,279 (17.1%) | 1,166 (15.6%) | 1,085 (7.0%) | 1,008 (6.5%) |
| Solid cancer | 5 (0.2%) | <5 (<5%) | 209 (0.6%) | 161 (0.4%) | 30 (0.4%) | 17 (0.2%) | 134 (0.9%) | 109 (0.7%) |
| Blood cancer | 0 (0.0%) | 0 (0.0%) | 44 (0.1%) | 27 (0.1%) | <5 (<5%) | <5 (<5%) | 10 (0.1%) | 10 (0.1%) |
| Immunodeficiency | 0 (0.0%) | <5 (<5%) | 26 (0.1%) | 27 (0.1%) | 15 (0.2%) | 8 (0.1%) | 60 (0.4%) | 45 (0.3%) |
| Prior vaccinations*, N (%) |  |  |  |  |  |  |  |  |
| Tdap | 1,562 (57.0%) | 1,439 (52.5%) | - | - | 3,329 (44.5%) | 3,366 (45.0%) | 90 (0.6%) | 97 (0.6%) |
| Influenza | 964 (35.2%) | 830 (30.3%) | - | - | 2,107 (28.2%) | 1,752 (23.4%) | 4,307 (27.9%) | 3,154 (20.4%) |

COPD: Chronic obstructive lung disease; CPRD: Clinical Practice Research Datalink, UK; SCIFI- PEARL: Swedish Covid-19 Investigation for Future Insights – a Population Epidemiology Approach using Register Linkage, Sweden; SIDIAP: Information System for Research in Primary Care, Spain; Tdap: tetanus, diphtheria and pertussis; UiO: University of Oslo, Norway.
"Exposed" refers to pregnant women who received the first dose, while "unexposed" refers to unvaccinated pregnant women. Matching was performed on a 1:1 basis using weekly sequential matching. Index date for the matched pair was defined as the date of the first dose vaccination.
\* Assessed in the year prior. Data on Tdap and influenza vaccines were not available in SCIFI-PEARL.
\*\* Assessed any time prior to index date, except for solid and blood cancers (assessed in the 5 years prior).

**Table 2.** Baseline characteristics of matched pregnant women included for the analysis of VE booster (3^rd^ dose) vs. primary (2-dose) vaccination

|  | **CPRD GOLD** |  | **SCIFI-PEARL** |  | **SIDIAP** |  | **UiO** |  |
| --- | --- | --- | --- | --- | --- | --- | --- | --- |
| Variable name | Exposed | Matched unexposed | Exposed | Matched unexposed | Exposed | Matched unexposed | Exposed | Matched unexposed |
| Number records, N | 1,892 | 1,892 | 26,195 | 26,195 | 2,218 | 2,218 | 10,791 | 10,791 |
| Age, Median [Q25 - Q75] | 32 [29-35] | 32 [29- 35] | 31 [29 - 34] | 31 [29 - 34] | 34 [31-37] | 34 [31- 37] | 31 [28-34] | 31 [28 - 34] |
| Vaccine brand, N (%) |  |  |  |  |  |  |  |  |
| BNT162b2 | 1,224 (64.7%) | - | 21,187 (80.9%) | - | 673 (30.3%) | - | 8,771 (81.3%) | - |
| mRNA-1273 | 668 (35.3%) | - | 5,008 (19.1%) | - | 1,545 (69.7%) | - | 2,020 (18.7%) | - |
| Trimester at index date, N (%) |  |  |  |  |  |  |  |  |
| Trimester 1 | 894 (47.3%) | 894 (47.3%) | 5,558 (21.2%) | 5,558 (21.2%) | 719 (32.4%) | 719 (32.4%) | 3,372 (31.2%) | 3,372 (31.2%) |
| Trimester 2 | 712 (37.6%) | 712 (37.6%) | 15,411 (58.8%) | 15,411 (58.8%) | 1,018 (45.9%) | 1,018 (45.9%) | 6,200 (57.5%) | 6,200 (57.5%) |
| Trimester 3 | 286 (15.1%) | 286 (15.1%) | 5,226 (20.0%) | 5,226 (20.0%) | 481 (21.7%) | 481 (21.7%) | 1,219 (11.3%) | 1,219 (11.3%) |
| Health visits*, Median [Q25 - Q75] | 30 [18-46] | 28 [16 - 44] | 6 [0 - 20] | 4 [0- 15] | 30 [16 - 52] | 26 [14 - 46] | 26 [16-44] | 24 [12 - 40] |
| Conditions**, N (%) |  |  |  |  |  |  |  |  |
| Asthma | 145 (7.7%) | 131 (6.9%) | 1,310 (5.0%) | 1,081 (4.1%) | 145 (6.5%) | 133 (6.0%) | 1,700 (15.8%) | 1,589 (14.7%) |
| Chronic hypertension | 25 (1.3%) | 30 (1.6%) | 870 (3.3%) | 895 (3.4%) | 53 (2.4%) | 35 (1.6%) | 816 (7.6%) | 806 (7.5%) |
| COPD | 0 (0.0%) | 0 (0.0%) | 9 (0.0%) | 6 (0.0%) | <5 (<5%) | <5 (<5%) | 76 (0.7%) | 67 (0.6%) |
| Diabetes | 70 (3.7%) | 61 (3.2%) | 627 (2.4%) | 596 (2.3%) | 111 (5.0%) | 91 (4.1%) | 519 (4.8%) | 536 (5.0%) |
| Obesity | 236 (12.5%) | 228 (12.1%) | 1,598 (6.1%) | 1,541 (5.9%) | 345 (15.6%) | 289 (13.0%) | 861 (8.0%) | 836 (7.7%) |
| Solid cancer | <5 (<5%) | <5 (<5%) | 182 (0.7%) | 105 (0.4%) | 10 (0.5%) | <5 (<5%) | 106 (1.0%) | 80 (0.7%) |
| Blood cancer | 0 (0.0%) | 0 (0.0%) | 15 (0.1%) | 6 (0.0%) | 0 (0.0%) | 0 (0.0%) | 19 (0.2%) | 7 (0.1%) |
| Immunodeficiency | 0 (0.0%) | <5 (<5%) | 24 (0.1%) | 16 (0.1%) | <5 (<5%) | <5 (<5%) | 44 (0.4%) | 25 (0.2%) |
| Prior vaccinations*, N (%) |  |  |  |  |  |  |  |  |
| Tdap | 995 (52.6%) | 1,000 (52.9%) | - | - | 887 (40.0%) | 934 (42.1%) | 6,534 (60.6%) | 84 (0.8%) |
| Influenza | 665 (35.1%) | 655 (34.6%) | - | - | 1,228 (55.4%) | 1,063 (47.9%) | 69 (0.6%) | 5,481 (50.8%) |

COPD: Chronic obstructive lung disease; CPRD: Clinical Practice Research Datalink, UK; SCIFI- PEARL: Swedish Covid-19 Investigation for Future Insights – a Population Epidemiology Approach using Register Linkage, Sweden; SIDIAP: Information System for Research in Primary Care, Spain; Tdap: tetanus, diphtheria and pertussis; UiO: University of Oslo, Norway.
"Exposed" refers to pregnant women who received a 1<sup>st</sup> booster (3<sup>rd</sup> dose), while "unexposed" refers to pregnant women who had received the second dose at least 3 months prior. Matching was performed on a 1:1 basis using weekly sequential matching. Index date for the matched pair was defined as the date of the third dose vaccination.
\* Assessed in the year prior. Data on Tdap and influenza vaccines were not available in SCIFI-PEARL.
\*\* Assessed any time prior to index date, except for solid and blood cancers (assessed in the 5 years prior).

Median follow-up of the matched pairs ranged from 42 [interquartile range, 25th-75th percentiles: 33-81] to 55 days [39-120] for the first trial (across databases), and from 73 days [18-172] to 116 [58-178] for the second trial. For the first trial, receiving a second dose more than 6 weeks after the first dose and thus deviating from the assigned treatment strategy was amongst the most frequent reason for censoring in CPRD GOLD in SCIFI-PEARL, affecting 70% and 35% of pregnancies included. Vaccination of the unexposed counterpart during follow up was also a common reason for censoring, affecting approximately 30-36% of pregnancies in SCIFI-PEARL, SIDIAP and UiO. For the second trial, pregnancy end was the main reason for censoring across databases (observed between 54% to 78% of pregnancies), followed by the unexposed counterpart receiving a booster during follow up (between 20% to 45% of pregnancies). Details on the numbers and reasons for censoring can be found in Supplementary Tables 11-18.

The number of patients matched in the study over calendar time can be found in Supplementary Figure 5. A peak in the uptake of the first dose was observed between May and September 2021, just preceding the time when the Delta variant became dominant (August to early December 2021). The uptake of the 1^st^ booster (3^rd^ dose) peaked between December 2021 and February 2022 across all databases, coinciding with the time period dominated by the Omicron variant. In both cases, uptake was somewhat faster in the UK compared to the other 3 contributing countries.

### Vaccine Effectiveness

For the first trial, VE of primary (2-dose) vaccination against infection compared to unvaccinated was estimated at 68.5% (95% CI 42.3 to 82.7) in CPRD GOLD, 25.0% (18.5 to 30.9) in SCIFI-PEARL,17.4% (7.8 to 25.9) in SIDIAP, and 27.7% (19.3 to 35.2) in UiO. VE against hospital admission was estimated at 53.6% (35.7 to 66.5) in SCIFI-PEARL, 57.5% (27.2 to 75.1) in SIDIAP and 57.7% (35.8 to 72.1) in UiO. Meta-analysis showed that VE for primary vaccination was 26.2% (16.2–35.1; I² = 0.7) against COVID-19 infection and 55.6% (44.1–64.8; I² = 0) against COVID-19 related hospitalisation (Figure 2).

**Figure 2.**
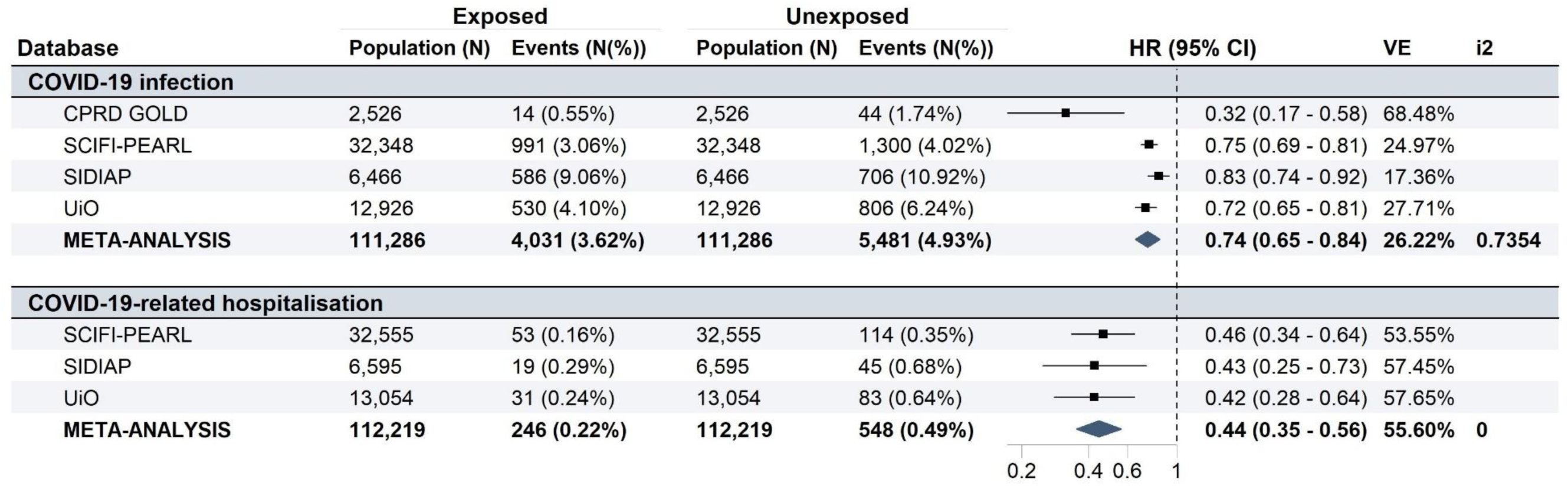
Vaccine effectiveness estimates: Primary (2-dose) vaccination vs. unvaccinated. Meta-analytic estimates of VE were derived from random-effects meta-analysis. CI: Confidence Interval; CPRD: Clinical Practice Research Datalink, UK; HR: Hazard Ratio; SCIFI-PEARL: Swedish Covid-19 Investigation for Future Insights – a Population Epidemiology Approach using Register Linkage, Sweden; SIDIAP: Information System for Research in Primary Care, Spain; UiO: University of Oslo, Norway; VE: Vaccine Effectiveness.

For the second trial, VE of a 1^st^ booster (3^rd^ dose) compared to primary (2-dose) vaccination against COVID-19 during pregnancy was estimated at 32.9% (11.2 to 49.2) in CPRD GOLD, 21.1% (8.6 to 31.8) in SIDIAP, 13.3% (4.8 to 21.1) in SCIFI-PEARL and 57.5% (54.2 to 60.6) in UiO. VE against hospitalisation was 52.4% (27.4 to 68.7) in SCIFI-PEARL and 32.8% (3.7 to 53.1) in UiO. VE against hospitalisation in SIDIAP had wide CI (-18.0% [-155.2 to 45.4]) due to the low number of events captured (unexposed: 12 vs. exposed:14). Meta-analysis showed that VE for a 1^st^ booster (3^rd^ dose) was 33.6% (-1.6 to 56.6; I^2^ =1) against COVID-19 infection and 34.1% (0.6 to 56.4; I^2^ =0.5) against COVID-19 related hospitalisation (Figure 3).

**Figure 3.**
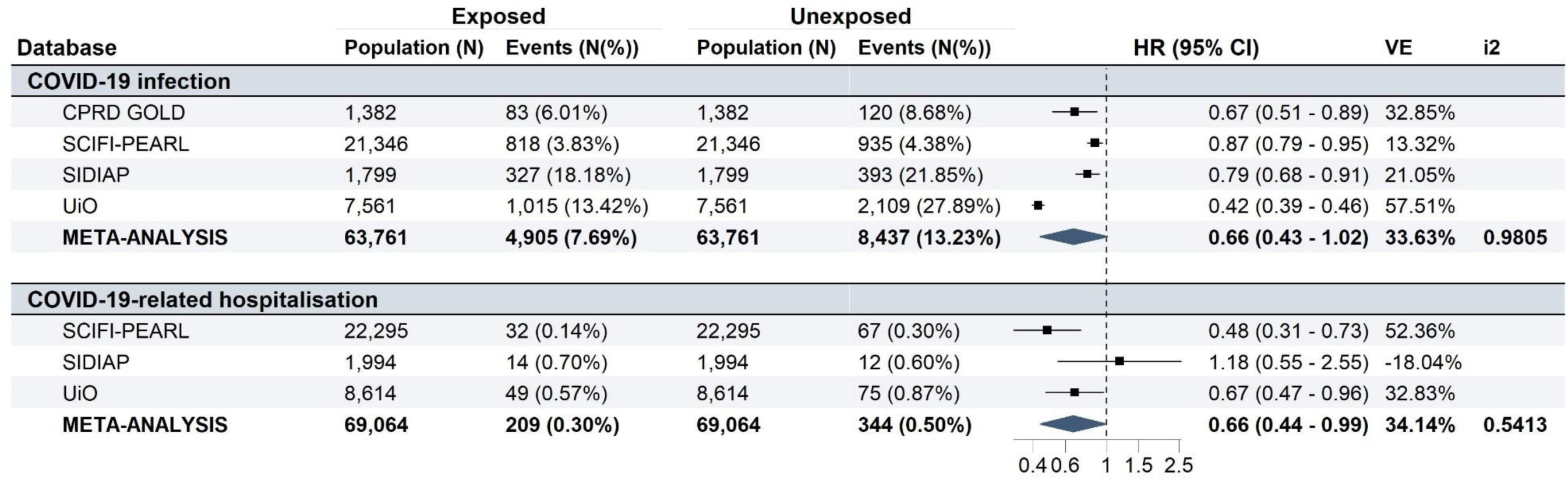
Vaccine effectiveness estimates: 1^st^ booster (3^rd^ dose) vs. primary (2-dose) vaccination. Meta-analytic estimates of VE were derived from random-effects meta-analysis. CI: Confidence Interval; CPRD: Clinical Practice Research Datalink, UK; HR: Hazard Ratio; SCIFI-PEARL: Swedish Covid-19 Investigation for Future Insights – a Population Epidemiology Approach using Register Linkage, Sweden; SIDIAP: Information System for Research in Primary Care, Spain; UiO: University of Oslo, Norway; VE: Vaccine Effectiveness.

We could not evaluate VE against ICU admission in either of the two trials, due to insufficient counts across groups compared. No significant differences in VE against COVID-19 infection or hospitalisation were observed across trimesters except for CPRD GOLD (Figure 4). Results stratified by vaccine brand showed similar estimates (Supplementary Figure 6). Kaplan-Meier curves and Log-Log survival curves suggested waning effectiveness over time (Supplementary Figures 7-10). HRs against COVID-19 infection using different time windows are reported in Supplementary Figure 11. NCOs with sufficient counts for assessment ranged between 20-33 NCOs across databases, except for CPRD GOLD (7-9 NCOs, depending on the trial). Among those with sufficient counts, <15% of NCOs were associated with vaccination and were not suggestive of residual confounding (Supplementary Figures 12-15). Results from the secondary analysis (up to end of data availability) were consistent with the main analysis (Supplementary Figures 16-19).

**Figure 4:**
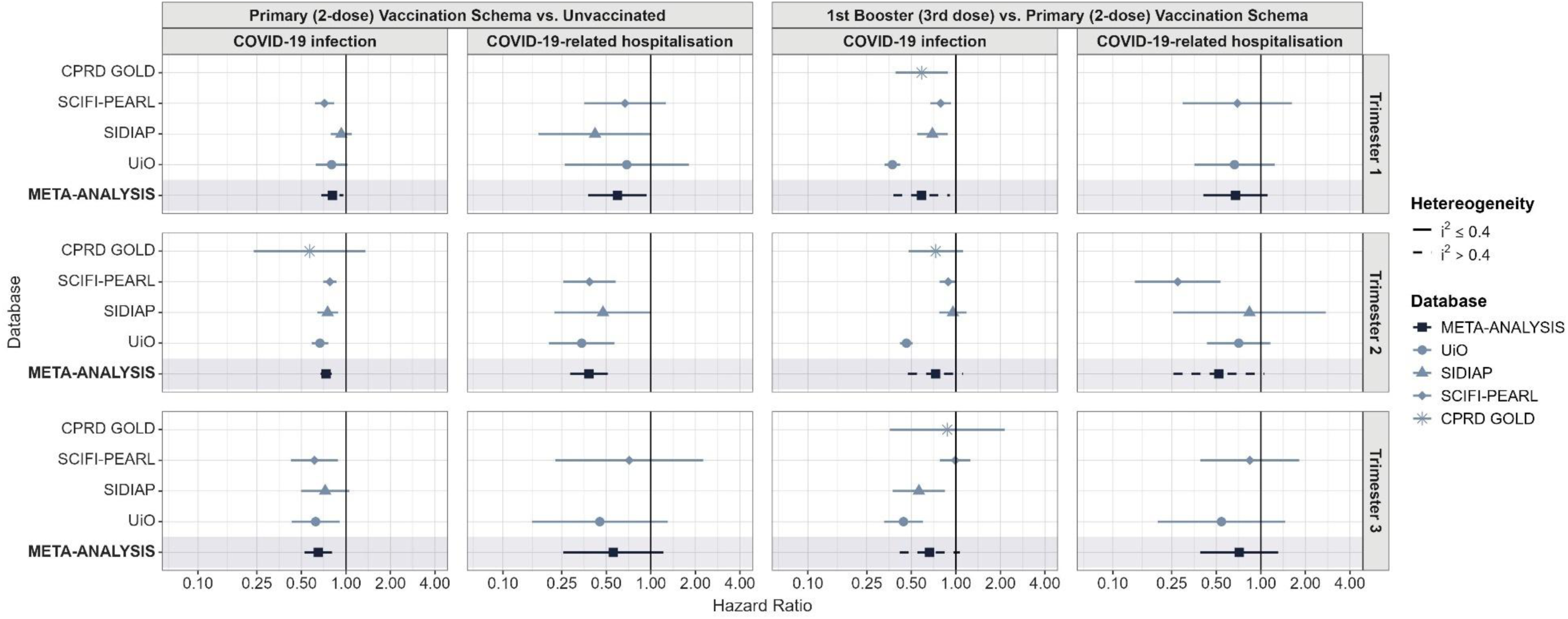
Hazard Ratios for COVID-19 Infection and Hospitalisatiom stratified by pregnancy trimester at time of vaccination: Primary (2-dose) Vaccination Schema vs. Unvaccinated, and Booster (3^rd^ dose) vs. Primary (2-dose) Vaccination Schema. Meta-analytic estimates of VE were derived from random-effects meta-analysis. CPRD: Clinical Practice Research Datalink, UK; HR: Hazard Ratio; SCIFI-PEARL: Swedish Covid-19 Investigation for Future Insights – a Population Epidemiology Approach using Register Linkage, Sweden; SIDIAP: Information System for Research in Primary Care, Spain; UiO: University of Oslo, Norway

### Sensitivity analyses

VE against COVID-19-related hospitalisation was reduced when including hospital admissions related to delivery, decreasing from 55.6% (44.1, 64.8 to I^2^ =0.6) to 28.2% (20.9 to 34.8; I^2^ =0.1) for the first trial, and 34.1% (0.6 to 56.4; I^2^ = 0.3) to 19.5% (4.2 to 32.4; I^2^ =0.3) for the second trial (Supplementary Figure 20). When restricting the definition of infection to those with a recorded positive test result, VE against COVID-19 was similar to that obtained when considering positive test results and compatible clinical codes. However, some estimates for VE for the 1^st^ booster (3^rd^ dose) were altered, especially in UiO, where the VE estimate against infection increased from 57.5% (54.2 to 60.1) to 78.3% (75.8 to 80.5) (Supplementary Figures 21-22).

## DISCUSSION

The findings of this multinational target trial emulation study including over 124,122 pregnancies suggest that primary (2-dose) vaccination against COVID-19 during pregnancy is effective in preventing infection during pregnancy with a VE of 26%, and a VE against COVID-19 related hospitalisations of 55%. Notably, a 1^st^ booster (3^rd^ dose) vaccination during pregnancy provided a 30-35% protection against COVID-19 infection and COVID-19 related hospitalisation compared to a primary (2-dose) vaccination. We could not evaluate VE against ICU admission due to the rarity of this outcome over the time periods studied.

Our results demonstrate lower levels of VE against both infection and hospitalisation compared to previous published literature. Prior pregnancy-specific literature has been mostly derived from Israeli health data, reporting short-term estimates for BNT162b2 vaccine only.(29) Dagan et al. estimated a VE of 96% (95% Cl 89 to 100) against infection and 89% (43 to 100) against COVID-19-related hospitalisation among pregnant women in days 7 to 56 after vaccination with a second dose during a time period dominated by the original SARS-CoV-2 reference strain and the B.1.1.7 (Alpha) variant. Regarding booster doses, Guedalia et al. estimated the effectiveness of a third vaccine given during pregnancy at least 5 months after the second dose compared to non-boosted pregnant women, reporting substantial differences in VE in time periods dominated by different SARS-CoV-2 variants. VE against hospitalisation was estimated at 92% (83 to 96) during a period dominated by the Delta variant, declining to 48% (37 to 57) when calculated during a period dominated by the Omicron variant. Discrepancies between our findings and those reported in prior studies may be attributable to different study periods, length of follow-up, settings and design choices. First, studies were conducted using data from time periods with different circulating SARS-CoV-2 variants. In our study, the main circulating variants when outcomes were observed were Delta and Omicron, with the latter being known to evade vaccine-induced immunity against infection.(15, 17, 30) When comparing results against the same variant, VE of a third dose estimated by Guedalia et. al during the Omicron period (48% [37 to 57]) was numerically higher than the meta-analytic estimate obtained in our study (34.1% [0.6 to 56.4]; I^2^ =0.5). Second, the strategies used to account for confounding differed across studies. Dagan et al. used exact matching on a subset of demographic and clinical variables, while Guedalia et. al only controlled for maternal age and parity. Our study used state-of-the-art target trial emulation methods, and included a more comprehensive approach to balance the groups and minimise confounding, including a much larger covariate set in the propensity scores. We also explicitly assessed covariate balance for a large set of conditions and medications using SMDs, and we used NCOs to test for remaining unobserved confounding. In addition, prior studies excluded individuals with previous documented COVID-19 infection,(16, 17, 31) whereas our study allowed these individuals to be included after a 90-days washout, while accounting for differences in testing. This approach likely reflects the current context, where a large majority of individuals have gained natural immunity through prior SARS-CoV-2 infection. Third, the median follow-up of matched participants in our study spanned from 42 to 55 days for the first trial and 73 to 116 days for the second trial, with the latter being slightly longer than that reported in prior studies.(16, 31) The rapid pace of vaccination campaigns caused many unexposed women to receive vaccination shortly after index date, which contributed to the censoring of data and shorter follow up. This was particularly relevant for the first trial, as many exposed individuals were censored due to not receiving the second vaccine dose within 6 weeks after the first dose as proscribed by their assigned treatment strategy. As a result, the estimates for primary (2-dose) vaccination in this study are likely to reflect the effectiveness of initiating vaccination instead of the effect of receiving the 2-dose regimen, particularly in CPRD GOLD and SCIFI-PEARL.

Comparisons of VE estimates derived from the emulation of the two target trials should be interpreted with caution, as they may be influenced by changes in testing practices and differences in case detection over time, which might have introduced differential misclassification of the outcome. Periods of more intensive testing efforts may have led to increased identification of asymptomatic or mild cases, potentially resulting in an underestimation of VE against infection. Our results also captured the effect of vaccine waning, which has been described in other studies.(30, 32, 33) We found a protective effect during the first 14 days after first-dose vaccination, which is unexpected in the days following vaccination given that immunity is gradually building.(34, 35) This has also been reported in other observational studies assessing COVID-19 VE, and might stem from differences in personal behavior during the period immediately before and after vaccinations.(27, 36)

Heterogeneity in VE was observed across databases, and therefore, results derived from the random-effects meta-analysis should be interpreted cautiously, especially in cases where the I^2^ statistic was suggestive of substantial heterogeneity (I^2^ ≥ 0.5).(37) Compared to other databases, VE against infection was significantly higher in CPRD GOLD in the first trial (68.5% [42.3, 82.7]), and in UiO in the second trial (57.5% [54.2, 60.6]). The heterogeneity observed is likely to be attributed to geographical differences in the timing and magnitude of pandemic waves, varying levels of herd immunity, and differences in national COVID-19 vaccination policies including dosage intervals, and timing for booster doses. Notably, a longer separation between the two primary doses was recommended in the UK (12 weeks) compared to other countries (typically 3 weeks) to increase vaccine coverage.(38) Some studies have suggested that the widening of this interval could lead to increased VE against COVID-19 infection,(39) potentially explaining the increased protection seen in CPRD GOLD compared to other data sources.

Our findings support that COVID-19 mRNA vaccination effectively prevents COVID-19-related hospitalisations during pregnancy, with a 1^st^ booster (3^rd^ dose) conferring additional substantial protection. These findings reinforce the effectiveness of COVID-19 vaccination and boosting during pregnancy to prevent severe COVID-19 outcomes in this population.

### Strengths & limitations

Our study has relevant strengths. To the best of our knowledge, this is the largest study on effectiveness of COVID-19 vaccines in pregnancy and the first study to provide results from different countries using a federated approach, improving the representativeness and geographical diversity of current studies on this topic. In addition, since most pregnancy-specific evidence has focused on the BNT162b2 vaccine, this study extends current knowledge by providing real-world effectiveness data for the mRNA-1273 vaccine.

Nevertheless, several limitations need to be considered. First, as in any observational analysis, assignment to treatment strategies was not randomised, and therefore, we cannot completely rule out residual confounding. We performed matching on a wide range of factors that may be expected to confound the causal effect of the vaccine on the studied outcomes and included NCOs to detect unmeasured confounding. However, we found an unlikely protective effect during the first 14 days after first-dose vaccination, which might be indicative of confounding due to unmeasured behavioural factors. Second, we only considered adjustment for baseline covariates under the assumption of no residual time-varying confounding after baseline, given the short median follow-up time of matched pairs. Third, we defined primary vaccination as two doses administered within a maximum of 42 or 49 days after a first dose vaccination with BNT162b2 or mRNA-1273, respectively. This led to many individuals, particularly in CPRD GOLD and SCIFI-PEARL, being censored. Further analysis with less restrictive intervals might provide a better estimation of VE in these settings. Fourth, low event counts prevented the assessment of VE against ICU admission and resulted in wide CI in some cases, especially when providing results stratified by pregnancy trimester or vaccine brand. Lastly, we did not study the effect of heterogenous vaccine regimens or schedules between vaccine doses, which were beyond the scope of this study and require further investigation.

## Conclusion

Primary (2-dose) vaccination showed 26.2% effectiveness against infection and 55.6% against hospitalisation, while a 1^st^ booster (3^rd^ dose) vaccination provided 33.6% effectiveness against infection and 34.1% against hospitalisation, indicating modest protection against infection.

## Acknowledgements

We acknowledge the significant scientific contributions of Professor Angela Lupattelli, who passed away during the course of this study. Professor. Lupattelli played an important role in the study’s design and contributed substantially to the handling of Norwegian data. Her dedication, insight, and expertise are highly appreciated. The authors thank Dr Carlen Reyes, who contributed to the identification of NCOs, Dr Alicia Abellan and colleagues, who developed and implemented the OMOP pregnancy extension table, which was instrumental in the development of this study, and Dr Marissa Erin Leblanc for her valuable suggestions.

## Declaration of interests

All authors have completed the ICMJE uniform disclosure form at https://www.icmje.org/disclosure-of-interest/ and declare no support from any organisation for the submitted work; no financial relationships with any organisations that might have an interest in the submitted work in the previous three years; no other relationships or activities that could appear to have influenced the submitted work. FN reports research project support (paid to his institution, no personal fees) by Bayer, with no relation to the work reported here. FN owns some AstraZeneca shares. TB reports consultancy for Institut Biochimique SA, with no relation to the work reported here.

## Funding

BR and TD-S work for a research group that receives/received unconditional research grants from UCB, and Johnson and Johnson, Innovative Medicines Initiative, European Medicines Agency, none of which relate the content of this manuscript. DPA’s research group from the University of Oxford has received research grants from the European Medicines Agency, from the Innovative Medicines Initiative, from Gilead Science, and from UCB Biopharma, none of which related to this manuscript.

FN has funding for this research from SciLifeLab / the Knut and Alice Wallenberg Foundation (KAW 2021-0010/ VC2021.0018 and KAW 2020.0299/VC 2022.0008) and the Swedish Research Council (2021-05045 and 2021-05450). The overarching SCIFI-PEARL study (contributing Swedish data to the current analysis) through P.I. FN also has core funding from the Swedish state under the agreement between the Swedish government and the county councils, the ALF-agreement (Avtal om Läkarutbildning och Forskning/Medical Training and Research Agreement), grants ALFGBG-938453, ALFGBG-971130, ALFGBG-978954, ALFGBG-1006729, a grant from FORTE (Research Council for Health, Working Life, and Welfare), grant 2024-01711, and previously from a joint grant from FORTE and FORMAS (Research Council for Environment, Agricultural Sciences and Spatial Planning), grant 2020-02828. Data acquisition of the Norwegian data in this study was funded by Professor Nordeng’s EU-COVID-19 project, funded by the Norwegian Research Council’s COVID-19 Emergency Call (project no. 31270).

## Ethical approval

This project was approved by the Clinical Research Ethics Committee of the IDIAPJGol (project code: 23/023-EOm), the CPRD’s Research Data Governance Process (Protocol No 21_000557), the Regional Committee for Research Ethics (approval number: 155294 /REK Nord), the Data Protection Officer at the University of Oslo (approval number: 523275) and the Swedish Ethical Review Authority (EPM 2020-01800 with subsequent amendments).

## Data availability statement

Patient level data cannot be shared without approval from data custodians owing to local information governance and data protection regulations. Aggregated data, analytical code, and detailed definitions of algorithms for identifying the events are available in a GitHub repository (https://github.com/rwepi-idiapjgol/COVIDVaccineEffectivenessDuringPregnancy). The results are available in an interactive web app (https://dpa-pde-oxford.shinyapps.io/COVID19VaccineEffectivenessDuringPregnancy/).

## Contributors

NMB and BR are joint first authors. NMB, BR, TD-S and HMEN conceived the study. NT, AL, HL, TB, DM and FN contributed to the study design. HMEN, NT, SH, FI, HL, TD-S, AG, TB, DPA and NMB contributed to the study execution (data holders). NMB and BR wrote the first manuscript draft, with valuable input from DPA. All coauthors interpreted the results and contributed to revision of the manuscript. All authors approved the final version and had final responsibility for the decision to submit for publication. TD-S and HMEN are guarantors. The corresponding author attests that all listed authors meet authorship criteria and that no others meeting the criteria have been omitted.

## Transparency

The corresponding author (the manuscript’s guarantor) affirms that the manuscript is an honest, accurate, and transparent account of the study being reported; that no important aspects of the study have been omitted; and that any discrepancies from the study as planned (and, if relevant, registered) have been explained.

## Notes

### Funding Statement

This study did not receive any funding

### Author Declarations

This project was approved by the Clinical Research Ethics Committee of the Primary Health Care Research Jordi Gol i Gurina (IDIAPJGol) (project code: 23/023-EOm), the Clinical Practice Research Datalink (CPRD) Research Data Governance Process (Protocol No 21_000557), the Regional Committee for Research Ethics (approval number: 155294 /REK Nord), the Data Protection Officer at the University of Oslo (approval number: 523275) and the Swedish Ethical Review Authority (EPM 2020-01800 with subsequent amendments).

