## Supplementary Material for "Effectiveness of COVID-19 mRNA primary and booster vaccination against infection and hospitalisation during pregnancy: a target trial emulation and meta-analysis of data from 4 European countries"

**Supplementary Table 1:** Specifications of the target trials.

| Protocol component | Target trial specification | Target trial emulation |
|---|---|---|
| **Eligibility criteria** | • Enrolment period: From the start of the vaccination campaign to 90 days prior to end of data availability<br>• Pregnant population (gestational age < 34 weeks)<br>• Aged 12 to 55 years<br>• No prior COVID-19 infection in the 90 days prior<br>• Enrolled in the health care system of interest for at least 12 months prior to pregnancy start. | Same as for the target trial |
| **Treatment strategies** | <u>Primary vaccination vs. unvaccination:</u><br>1. Primary vaccination schema (2 doses) initiated during pregnancy:<br>a. BNT162b2 (gap between doses 17 to 42 days)<br>b. mRNA-1273 (gap between doses 24 to 49 days)<br>2. No vaccination<br><u>Booster dose vs. primary vaccination:</u><br>1. Third dose of BNT162b2 or mRNA-1273 during pregnancy after at least 3 months after the 2n dose.<br>2. Primary vaccination | Same as for the target trial |
| **Treatment assignment** | Individuals are randomly assigned to a strategy at baseline and will be aware of the strategy to which they have been assigned | Individuals are classified according to the strategy with which their data are compatible at baseline. Randomisation is emulated by adjusting for baseline confounders. |
| **Outcomes** | • SARS-CoV-2 infection<br>• COVID-19-related hospitalisation<br>• COVID-19-related ICU admission | Same as for the target trial |
| **Follow-up** | • Starts at treatment assignment<br>• Ends at the first of: outcome occurrence, loss of follow-up, end of data availability or deviation from the treatment assigned at baseline. | Same as for the target trial |
| **Causal contrast** | Per-protocol effect | Same as for the target trial |
| **Statistical analyses** | Per-protocol analysis: censor participants when they deviate from their assigned treatment strategy. | Same as for the target trial |

**Supplementary Text 1:** Descriptions of the data sources.

This network study was conducted using four routine healthcare databases with complete vaccination coverage from the UK, Sweden, Spain and Norway. All databases were previously mapped to the Observational Medical Outcomes Partnership (OMOP) Common Data Model, which allowed a federated approach. Data cut for all participating databases was between 2023 and 2024 (CPRD GOLD: 23-06-2023; SCIFI-PEARL: 30-08-2024; SIDIAP: 30-06-2023; UiO: 31-12-2023).

Clinical Practice Research Datalink (CPRD) GOLD, UK:
The Clinical Practice Research Datalink (CPRD) is a governmental, not-for-profit research service, jointly funded by the National Institute for Health and Care Research and the Medicines and Healthcare products Regulatory Agency, a part of the Department of Health and Social Care (https://cprd.com). CPRD collects anonymised patient data from a network of GP practices across the UK.(1) It contains of all clinical and referral events in primary care in addition to comprehensive demographic information and medication prescription. Data from contributing practices are collected and processed into research databases. Data are available for 20 million patients. Pregnancy episodes in CPRD were inferred using a previously validated algorithm, which was validated for CPRD data mapped to the OMOP common data model.(2)

Swedish Covid-19 Investigation for Future Insights – a Population Epidemiology Approach using Register Linkage (SCIFI-PEARL), Sweden:
Swedish data was obtained through the Swedish Covid-19 Investigation for Future Insights – a Population Epidemiology Approach using Register Linkage (SCIFI-PEARL) project. The study population covers the full Swedish population since 2015 and links from multiples using the unique Swedish personal identification number. Data mapped to OMOP common data model include high-quality data on specialist outpatient care and inpatient care from the National Patient Register (3), intensive care from the Swedish Intensive Care Register (4), mortality data from the National Cause-of-death Register (5), medication information from the National Prescribed Drug Register (6), SARS-CoV-2 test positive records from the National monitoring system for communicable diseases (SmiNet) (7), COVID-19 vaccination data from the National Vaccination Register.(3-7) It also includes demographics from the Total Population Register, socioeconomic information from the Longitudinal integrated database for health insurance and labour market studies (LISA) and more.(8,9) Primary care data is available for 40% of patients only. Additionally, OMOP mapped data from the National Medical Birth Register is also included, with information on mother and child, with a national coverage of about 100,000 births per year.(10)

Information System for Research in Primary Care (SIDIAP), Spain:
The Information System for Research in Primary Care (SIDIAP; www.sidiap.org) database in Catalonia (SIDIAP) contains pseudo-anonymised electronic health records since 2006 for 8 million people (75% of the Catalan population) and is representative of the general population in terms of age, sex, and geographic distribution.(11) SIDIAP includes high-quality data registered by healthcare professionals on sociodemographic, disease diagnoses, anthropometric measurements, vaccine administration, prescription, and dispensation of drugs, among others. The database includes detailed pregnancy information such as dates of last period and of estimated delivery, along with the type of delivery, the circumstance of the end of the pregnancy (e.g. type of delivery, abortion, etc.), gestational age and trimestral obstetric ultrasounds, among others. SIDIAP has been linked to SARS-CoV-2 Reverse Transcription Polymerase Chain Reaction (RT-PCR) test results, hospital discharge records (CMBD, Conjunto Mínimo Básico de Datos), and regional mortality data.

Norwegian Linked Health Registries, Norway:
Data from Norway was obtained from the Norwegian Linked Health Registries at the University of Oslo (UiO), which contains data from Norwegian health care administrative databases that cover the entire Norwegian population from 2008 to 2023. The data that UiO mapped onto the OMOP CDM includes high-quality data

on disease diagnoses from primary and secondary care, drug dispensations, vaccine administration, positive tests for communicable diseases, demographic characteristics, among others from the Norwegian Prescribed Drug Register (LMR), the national vaccination registry (SYSVAK), the Norwegian Surveillance System for Communicable Diseases (MSIS) , Norwegian Control and Payment of Health Reimbursements Database (KUHR), and the Norwegian Patient Registry (NPR).(12-15) Importantly, it includes detailed information related to all pregnancies in Norway (information on the mother, father, and child), approximately 60,000 deliveries per year from the Medical Birth Registry of Norway (MBRN).(16) The MBRN contains all pregnancies with more than 12 week duration. For this study, we have used an algorithm developed by the UiO that allows the identification and dating of pregnancies during the first 12 weeks of gestation.(17)

<u>References:</u>

**Supplementary Figure 1:** Study Design Diagram and Propensity Score Covariates.

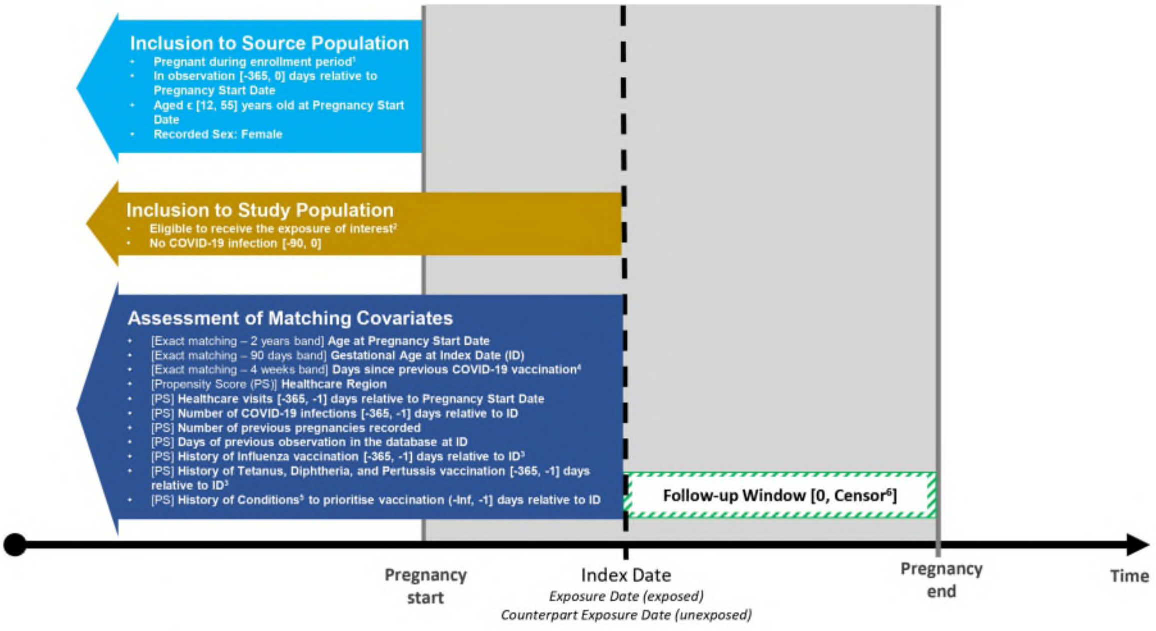

[1] Enrolment period: Start of the vaccination campaign (December 2020) to 90 days before end of available data

[2] Unvaccinated (Primary vaccination schema vs. Unvaccinated) before Index Date, and 90 days or more since 2nd dose at Index Date (Booster vs. Primary vaccination schema)

[3] Influenza and Tdap (Tetanus, Diphtheria, and Pertussis ) vaccination where not assessed for SCIFI-PEARL database as this information was not available

[4] Exact matching on days since previous vaccination in 30-days band – Only for Booster vs. Primary vaccination schema

[5] Conditions: Respiratory Conditions (asthma, chronic obstructive pulmonary disease, bronchiectasis, or bronchitis), Blood Cancer within the last 5 years, Cardiological Disease (excluding hypertension), Diabetes (any type), Immunodeficiency, Chronic Liver Disease, Obesity, Organ Transplant Recipient, and Solid Cancer within the past 5 years

[6] Follow-up end at first of: outcome of interest, pregnancy end date, end of data/death, or deviation from protocol strategy

**Supplementary Table 2:** Count and percentage of re-enrolled pregnant women across analyses and databases.

| Analysis | CDM name | N (%) |
|---|---|---|
| **1st 1st Booster (3rd dose) (3rd dose) vs. Primary (2-dose) Vaccination Schema** | CPRD GOLD | 333 (9.65%) |
| | SCIFI-PEARL | 7126 (15.74%) |
| | SIDIAP | 316 (7.67%) |
| | UiO | 2760 (14.66%) |
| **Primary (2-dose) Vaccination Schema vs. Unvaccinated** | CPRD GOLD | 561 (11.4%) |
| | SCIFI-PEARL | 10920 (17.65%) |
| | SIDIAP | 2205 (17.29%) |
| | UiO | 4907 (18.89%) |

**Supplementary Figure 2:** Flowchart for study populations in the CPRD GOLD database.

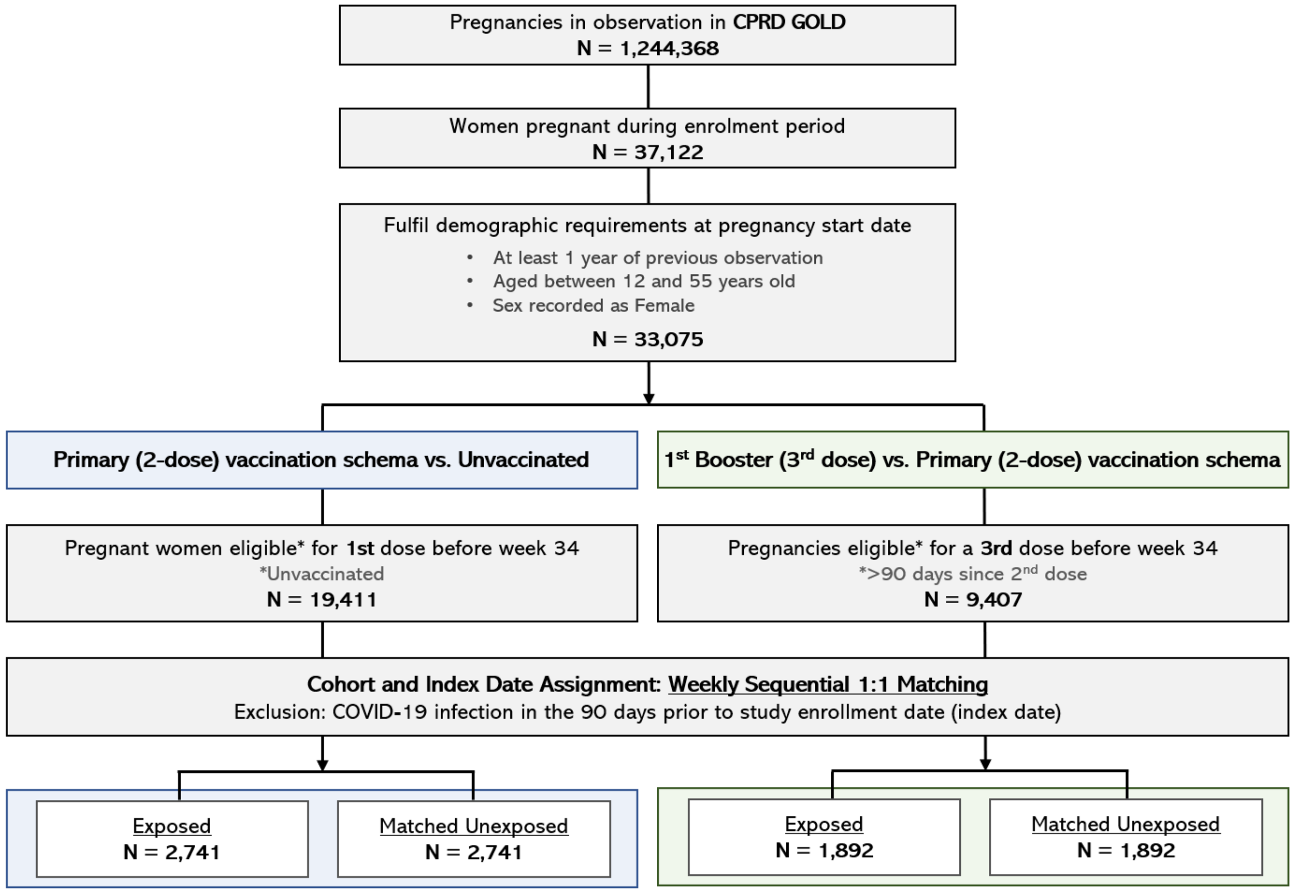

**Supplementary Figure 3:** Flowchart for study populations in the SIDIAP database.

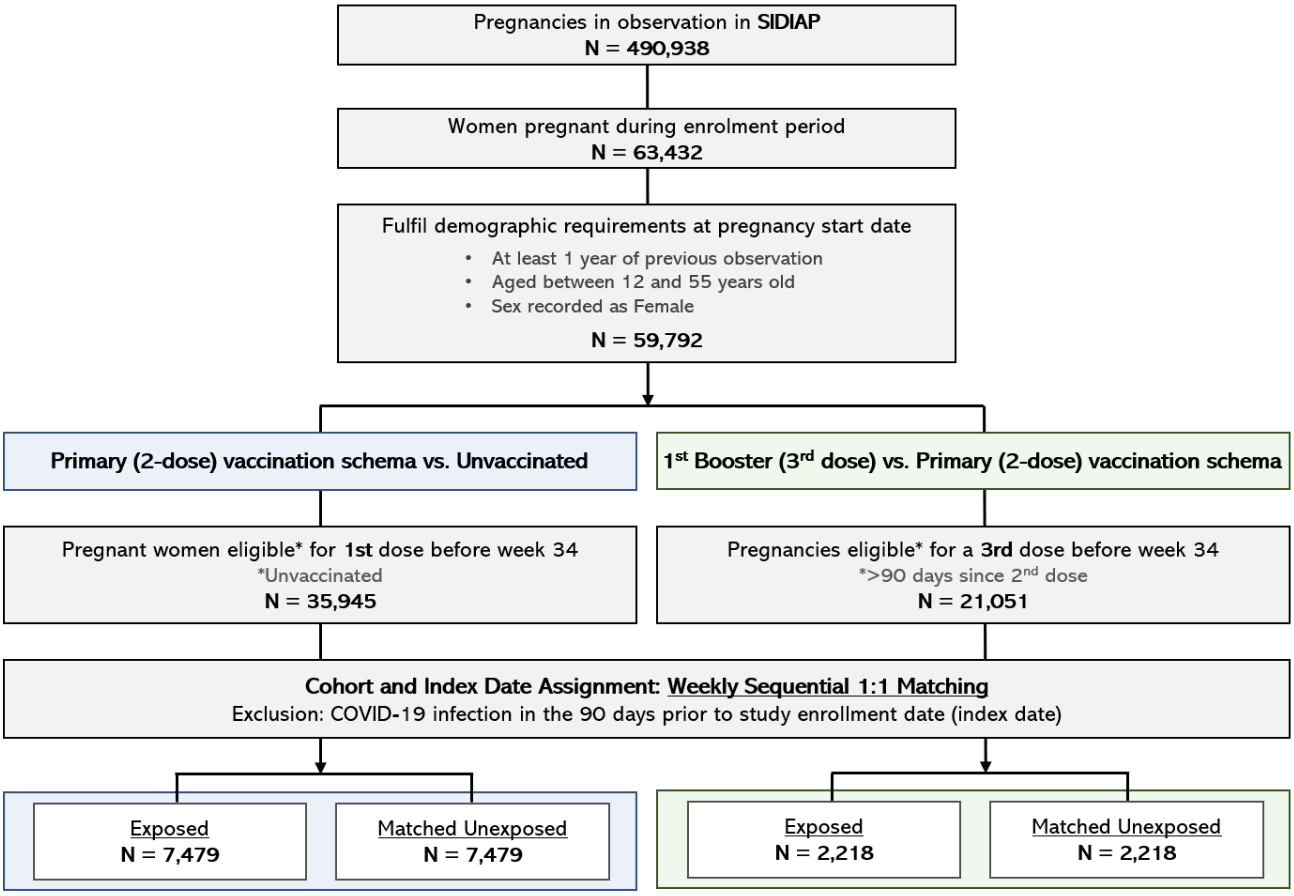

**Supplementary Figure 4:** Flowchart for study populations in the UiO database.

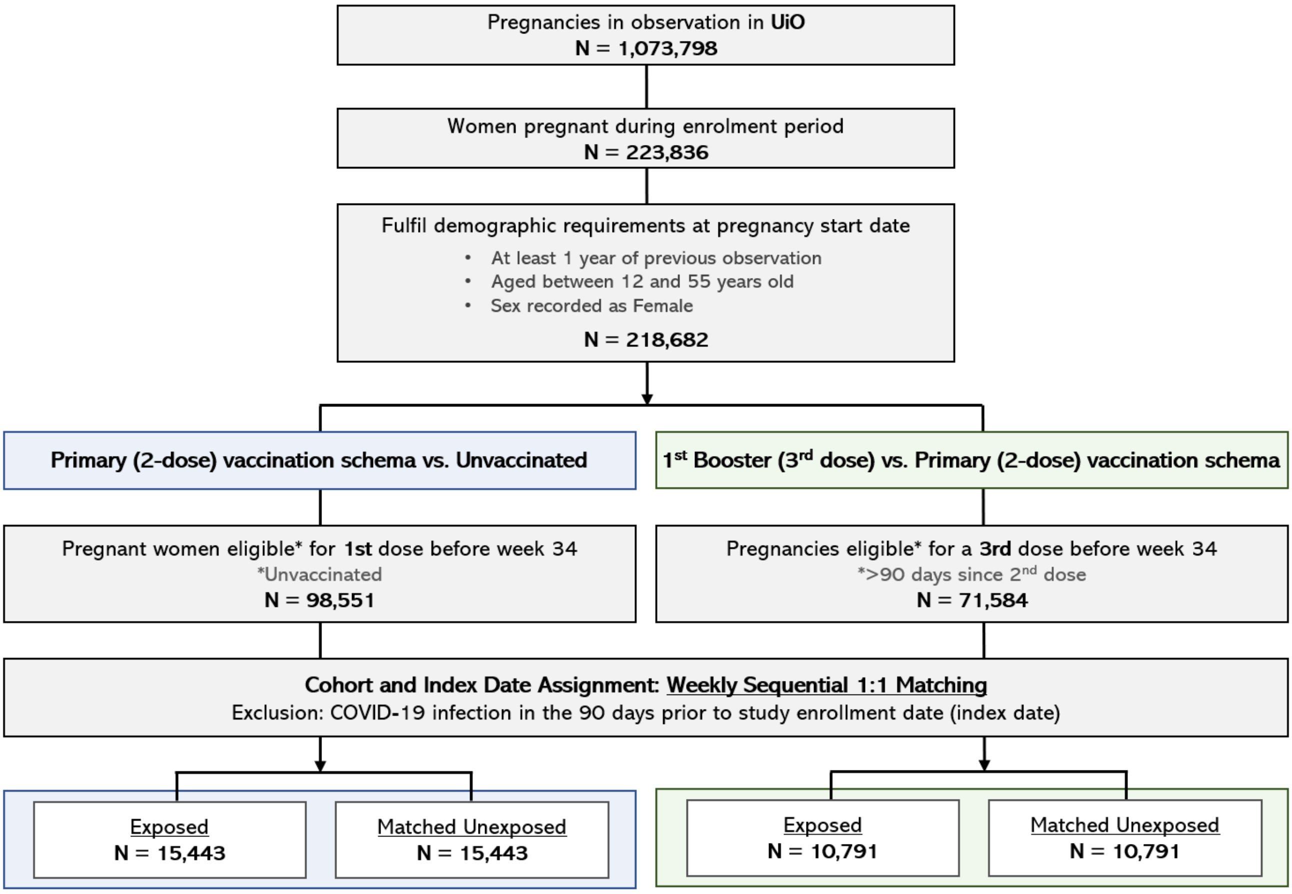

**Supplementary Table 3:** Baseline characteristics of the study population in the CPRD GOLD database, for the Primary (2-dose) Vaccination Schema vs. Unvaccinated analysis.

| Covariate | Covariate level | Estimate | Cohort | |
|---|---|---|---|---|
| | | | Unexposed | Exposed |
| **Number records** | - | N | 2,741 | 2,741 |
| **Age (Years)** | - | Median (Q25 - Q75) | 32.00 (28.00 - 35.00) | 32.00 (28.00 - 35.00) |
| **Age Group** | 12 to 24 | N (%) | 272 (9.9%) | 265 (9.7%) |
| | 25 to 39 | N (%) | 2,318 (84.6%) | 2,331 (85.0%) |
| | 40 to 55 | N (%) | 151 (5.5%) | 145 (5.3%) |
| **Gestational Trimester** | T1 | N (%) | 1,107 (40.4%) | 1,107 (40.4%) |
| | T2 | N (%) | 954 (34.8%) | 954 (34.8%) |
| | T3 | N (%) | 680 (24.8%) | 680 (24.8%) |
| **Vaccine Product** | Moderna | N (%) | - | 280 (10.2%) |
| | Pfizer | N (%) | - | 2,461 (89.8%) |
| **Previous Pregnancies** | - | Median (Q25 - Q75) | 2.00 (2.00 - 2.00) | 2.00 (2.00 - 2.00) |
| **Healthcare Visits (Past Year)** | - | Median (Q25 - Q75) | 20.00 (11.00 - 38.00) | 24.00 (14.00 - 42.00) |
| **Days of Prior Observation** | - | Median (Q25 - Q75) | 3,290 (1,394 - 6,545) | 3,020 (1,344 - 6,577) |
| **COVID-19 Infection (Any Time Prior)** | - | Median (Q25 - Q75) | 0.00 (0.00 - 0.00) | 0.00 (0.00 - 0.00) |
| **Other Vaccinations (Any Time Prior)** | Tdap | N (%) | 1,439 (52.5%) | 1,562 (57.0%) |
| | Influenza | N (%) | 830 (30.3%) | 964 (35.2%) |
| **Comorbidities (Any Time Prior)** | PCOS | N (%) | 64 (2.3%) | 62 (2.3%) |
| | Venous thromboembolism | N (%) | 11 (0.4%) | 7 (0.3%) |
| | Anemia | N (%) | 114 (4.2%) | 120 (4.4%) |

| Covariate | Covariate level | Estimate | Cohort | |
|---|---|---|---|---|
| | | | Unexposed | Exposed |
| | Pregnancy induced hypertension | N (%) | 12 (0.4%) | 20 (0.7%) |
| | Renal impairment | N (%) | 2 (0.1%) | 2 (0.1%) |
| | Stroke | N (%) | 2 (0.1%) | 0 (0.0%) |
| | Chronic hypertension | N (%) | 28 (1.0%) | 30 (1.1%) |
| | Heart conditions | N (%) | 12 (0.4%) | 23 (0.8%) |
| | Hypothyroidism | N (%) | 64 (2.3%) | 73 (2.7%) |
| | Anxiety disorder | N (%) | 229 (8.4%) | 185 (6.7%) |
| | Depressive disorder | N (%) | 444 (16.2%) | 372 (13.6%) |
| | Diabetes (any type) | N (%) | 94 (3.4%) | 110 (4.0%) |
| | Interstitial lung disease | N (%) | 0 (0.0%) | 1 (0.0%) |
| | Asthma | N (%) | 173 (6.3%) | 212 (7.7%) |
| | Pneumonia | N (%) | 12 (0.4%) | 12 (0.4%) |
| | HIV | N (%) | 0 (0.0%) | 0 (0.0%) |
| **Medications Prescribed (Last 180 Days)** | Estrogens | N (%) | 17 (0.6%) | 21 (0.8%) |
| | Drugs ulcer and gords | N (%) | 244 (8.9%) | 257 (9.4%) |
| | Antiinflammatory antirehumatic | N (%) | 151 (5.5%) | 136 (5.0%) |
| | Corticosteroids | N (%) | 20 (0.7%) | 30 (1.1%) |
| | Antithrombotics | N (%) | 33 (1.2%) | 33 (1.2%) |
| | Antihistamines systemic | N (%) | 422 (15.4%) | 436 (15.9%) |
| | Antidepressants | N (%) | 419 (15.3%) | 423 (15.4%) |
| | Progestogens | N (%) | 84 (3.1%) | 86 (3.1%) |
| | Antimycotics systemic | N (%) | 22 (0.8%) | 15 (0.5%) |
| | Immunosupressants | N (%) | 2 (0.1%) | 3 (0.1%) |
| | Antiepileptics | N (%) | 52 (1.9%) | 37 (1.3%) |
| | Antibacterials systemic | N (%) | 567 (20.7%) | 512 (18.7%) |

| Covariate | Covariate level | Estimate | Cohort | |
|---|---|---|---|---|
| | | | Unexposed | Exposed |
| | Analgesics and antipyretics | N (%) | 260 (9.5%) | 279 (10.2%) |

**Supplementary Table 4:** Baseline characteristics of the study population in the CPRD GOLD database, for the 1st 1st Booster (3rd dose) (3rd dose) vs. Primary (2-dose) Vaccination Schema analysis.

| Covariate | Covariate level | Estimate | Cohort | |
|---|---|---|---|---|
| | | | Unexposed | Exposed |
| **Number records** | - | N | 1,892 | 1,892 |
| **Age (Years)** | - | Median (Q25 - Q75) | 32.00 (29.00 - 35.00) | 32.00 (29.00 - 35.00) |
| **Age Group** | 12 to 24 | N (%) | 109 (5.8%) | 104 (5.5%) |
| | 25 to 39 | N (%) | 1,713 (90.5%) | 1,714 (90.6%) |
| | 40 to 55 | N (%) | 70 (3.7%) | 74 (3.9%) |
| **Gestational Trimester** | T1 | N (%) | 894 (47.3%) | 894 (47.3%) |
| | T2 | N (%) | 712 (37.6%) | 712 (37.6%) |
| | T3 | N (%) | 286 (15.1%) | 286 (15.1%) |
| **Vaccine Product** | Moderna | N (%) | - | 668 (35.3%) |
| | Pfizer | N (%) | - | 1,224 (64.7%) |
| **Previous Pregnancies** | - | Median (Q25 - Q75) | 2.00 (2.00 - 2.00) | 2.00 (2.00 - 2.00) |
| **Healthcare Visits (Past Year)** | - | Median (Q25 - Q75) | 28.00 (16.00 - 44.00) | 30.00 (18.00 - 46.00) |
| **Days of Prior Observation** | - | Median (Q25 - Q75) | 3,340 (1,441 - 6,708) | 2,768 (1,218 - 6,664) |
| **COVID-19 Infection (Any Time Prior)** | - | Median (Q25 - Q75) | 0.00 (0.00 - 0.00) | 0.00 (0.00 - 0.00) |
| **Other Vaccinations (Any Time Prior)** | Tdap | N (%) | 1,000 (52.9%) | 995 (52.6%) |
| | Influenza | N (%) | 655 (34.6%) | 665 (35.1%) |
| **Comorbidities (Any Time Prior)** | PCOS | N (%) | 49 (2.6%) | 59 (3.1%) |
| | Venous thromboembolism | N (%) | 11 (0.6%) | 8 (0.4%) |
| | Anemia | N (%) | 89 (4.7%) | 71 (3.8%) |
| | Pregnancy induced hypertension | N (%) | 12 (0.6%) | 12 (0.6%) |

| Covariate | Covariate level | Estimate | Cohort | |
|---|---|---|---|---|
| | | | Unexposed | Exposed |
| | Renal impairment | N (%) | 4 (0.2%) | 7 (0.4%) |
| | Stroke | N (%) | 1 (0.1%) | 2 (0.1%) |
| | Chronic hypertension | N (%) | 30 (1.6%) | 25 (1.3%) |
| | Heart conditions | N (%) | 19 (1.0%) | 10 (0.5%) |
| | Hypothyroidism | N (%) | 46 (2.4%) | 51 (2.7%) |
| | Anxiety disorder | N (%) | 124 (6.6%) | 110 (5.8%) |
| | Depressive disorder | N (%) | 293 (15.5%) | 228 (12.1%) |
| | Diabetes (any type) | N (%) | 61 (3.2%) | 70 (3.7%) |
| | Interstitial lung disease | N (%) | 0 (0.0%) | 0 (0.0%) |
| | Asthma | N (%) | 131 (6.9%) | 145 (7.7%) |
| | Pneumonia | N (%) | 6 (0.3%) | 8 (0.4%) |
| | HIV | N (%) | 0 (0.0%) | 0 (0.0%) |
| **Medications Prescribed (Last 180 Days)** | Estrogens | N (%) | 15 (0.8%) | 10 (0.5%) |
| | Drugs ulcer and gords | N (%) | 194 (10.3%) | 164 (8.7%) |
| | Antiinflammatory antirehumatic | N (%) | 109 (5.8%) | 78 (4.1%) |
| | Corticosteroids | N (%) | 32 (1.7%) | 28 (1.5%) |
| | Antithrombotics | N (%) | 21 (1.1%) | 27 (1.4%) |
| | Antihistamines systemic | N (%) | 351 (18.6%) | 297 (15.7%) |
| | Antidepressants | N (%) | 305 (16.1%) | 283 (15.0%) |
| | Progestogens | N (%) | 52 (2.7%) | 60 (3.2%) |
| | Antimycotics systemic | N (%) | 20 (1.1%) | 16 (0.8%) |
| | Immunosupressants | N (%) | 7 (0.4%) | 4 (0.2%) |
| | Antiepileptics | N (%) | 34 (1.8%) | 32 (1.7%) |
| | Antibacterials systemic | N (%) | 427 (22.6%) | 392 (20.7%) |
| | Analgesics and antipyretics | N (%) | 180 (9.5%) | 190 (10.0%) |

**Supplementary Table 5:** Baseline characteristics of the study population in the SCIFI-PEARL database, for the Primary (2-dose) Vaccination Schema vs. Unvaccinated analysis.

| Covariate | Covariate level | Estimate | Cohort | |
|---|---|---|---|---|
| | | | Unexposed | Exposed |
| **Number records** | - | N | 36,398 | 36,398 |
| **Age (Years)** | - | Median (Q25 - Q75) | 31 (28 - 35) | 31 (28 - 35) |
| **Age Group** | 12 to 24 | N (%) | 2,353 (6.5%) | 2,328 (6.4%) |
| | 25 to 39 | N (%) | 32,407 (89.0%) | 32,427 (89.1%) |
| | 40 to 55 | N (%) | 1,638 (4.5%) | 1,643 (4.5%) |
| **Gestational Trimester** | T1 | N (%) | 9,441 (25.9%) | 9,441 (25.9%) |
| | T2 | N (%) | 18,782 (51.6%) | 18,782 (51.6%) |
| | T3 | N (%) | 8,175 (22.5%) | 8,175 (22.5%) |
| **Vaccine Product** | Moderna | N (%) | - | 7,348 (20.2%) |
| | Pfizer | N (%) | - | 29,050 (79.8%) |
| **Previous Pregnancies** | - | Median (Q25 - Q75) | 1.00 (1.00 - 2.00) | 2.00 (2.00 - 2.00) |
| **Healthcare Visits (Past Year)** | - | Median (Q25 - Q75) | 4.00 (1.00 - 14.00) | 8.00 (2.00 - 22.00) |
| **Days of Prior Observation** | - | Median (Q25 - Q75) | 2,378 (2,338 - 2,413) | 2,379 (2,344 - 2,416) |
| **COVID-19 Infection (Any Time Prior)** | - | Median (Q25 - Q75) | 0.00 (0.00 - 0.00) | 0.00 (0.00 - 0.00) |
| **Comorbidities (Any Time Prior)** | Chronic hypertension | N (%) | 995 (2.7%) | 1,267 (3.5%) |
| | Heart conditions | N (%) | 388 (1.1%) | 474 (1.3%) |
| | Pneumonia | N (%) | 565 (1.6%) | 499 (1.4%) |
| | Venous thromboembolism | N (%) | 188 (0.5%) | 193 (0.5%) |

| Covariate | Covariate level | Estimate | Cohort | |
|---|---|---|---|---|
| | | | Unexposed | Exposed |
| | Anxiety disorder | N (%) | 4,483 (12.3%) | 4,908 (13.5%) |
| | Stroke | N (%) | 25 (0.1%) | 27 (0.1%) |
| | Anemia | N (%) | 3,535 (9.7%) | 2,978 (8.2%) |
| | Diabetes (any type) | N (%) | 463 (1.3%) | 563 (1.5%) |
| | Hypothyroidism | N (%) | 1,640 (4.5%) | 1,693 (4.7%) |
| | Asthma | N (%) | 1,502 (4.1%) | 1,778 (4.9%) |
| | PCOS | N (%) | 1,250 (3.4%) | 1,235 (3.4%) |
| | Depressive disorder | N (%) | 3,265 (9.0%) | 3,521 (9.7%) |
| | HIV | N (%) | 17 (0.0%) | 15 (0.0%) |
| | Interstitial lung disease | N (%) | 35 (0.1%) | 36 (0.1%) |
| | Renal impairment | N (%) | 57 (0.2%) | 77 (0.2%) |
| | Pregnancy induced hypertension | N (%) | 730 (2.0%) | 921 (2.5%) |
| **Medications Prescribed (Last 180 Days)** | Antithrombotics | N (%) | 590 (1.6%) | 620 (1.7%) |
| | Immunosupressants | N (%) | 167 (0.5%) | 234 (0.6%) |
| | Progestogens | N (%) | 2,605 (7.2%) | 2,917 (8.0%) |
| | Estrogens | N (%) | 1,461 (4.0%) | 1,711 (4.7%) |
| | Antimycotics systemic | N (%) | 466 (1.3%) | 443 (1.2%) |
| | Antidepressants | N (%) | 2,529 (6.9%) | 3,427 (9.4%) |
| | Analgesics and antipyretics | N (%) | 5,043 (13.9%) | 5,450 (15.0%) |
| | Corticosteroids | N (%) | 592 (1.6%) | 639 (1.8%) |
| | Antihistamines systemic | N (%) | 7,469 (20.5%) | 7,985 (21.9%) |
| | Antibacterials systemic | N (%) | 3,667 (10.1%) | 3,333 (9.2%) |
| | Drugs ulcer and gords | N (%) | 1,964 (5.4%) | 1,846 (5.1%) |
| | Antiepileptics | N (%) | 244 (0.7%) | 310 (0.9%) |

| Covariate | Covariate level | Estimate | Cohort | |
|---|---|---|---|---|
| | | | Unexposed | Exposed |
| | Antiinflammatory antirehumatic | N (%) | 1,337 (3.7%) | 1,326 (3.6%) |

**Supplementary Table 6:** Baseline characteristics of the study population in the SCIFI-PEARL database, for the 1st 1st Booster (3rd dose) (3rd dose) vs. Primary (2-dose) Vaccination Schema analysis.

| Covariate | Covariate level | Estimate | Cohort | |
|---|---|---|---|---|
| | | | Unexposed | Exposed |
| **Number records** | - | N | 26,195 | 26,195 |
| **Age (Years)** | - | Median (Q25 - Q75) | 31 (29 - 34) | 31 (29 - 34) |
| **Age Group** | 12 to 24 | N (%) | 1,121 (4.3%) | 1,085 (4.1%) |
| | 25 to 39 | N (%) | 24,203 (92.4%) | 24,244 (92.6%) |
| | 40 to 55 | N (%) | 871 (3.3%) | 866 (3.3%) |
| **Gestational Trimester** | T1 | N (%) | 5,558 (21.2%) | 5,558 (21.2%) |
| | T2 | N (%) | 15,411 (58.8%) | 15,411 (58.8%) |
| | T3 | N (%) | 5,226 (20.0%) | 5,226 (20.0%) |
| **Vaccine Product** | Moderna | N (%) | - | 5,008 (19.1%) |
| | Pfizer | N (%) | - | 21,187 (80.9%) |
| **Previous Pregnancies** | - | Median (Q25 - Q75) | 2.00 (1.00 - 2.00) | 2.00 (2.00 - 2.00) |
| **Healthcare Visits (Past Year)** | - | Median (Q25 - Q75) | 4.00 (0.00 - 15.00) | 6.00 (0.00 - 20.00) |
| **Days of Prior Observation** | - | Median (Q25 - Q75) | 2,591 (2,572 - 2,659) | 2,592 (2,574 - 2,665) |
| **COVID-19 Infection (Any Time Prior)** | - | Median (Q25 - Q75) | 0.00 (0.00 - 0.00) | 0.00 (0.00 - 0.00) |
| **Comorbidities (Any Time Prior)** | Chronic hypertension | N (%) | 895 (3.4%) | 870 (3.3%) |
| | Heart conditions | N (%) | 286 (1.1%) | 362 (1.4%) |
| | Pneumonia | N (%) | 370 (1.4%) | 366 (1.4%) |
| | Venous thromboembolism | N (%) | 123 (0.5%) | 131 (0.5%) |

| Covariate | Covariate level | Estimate | Cohort | |
|---|---|---|---|---|
| | | | Unexposed | Exposed |
| | Anxiety disorder | N (%) | 3,427 (13.1%) | 3,598 (13.7%) |
| | Stroke | N (%) | 18 (0.1%) | 19 (0.1%) |
| | Anemia | N (%) | 2,156 (8.2%) | 1,922 (7.3%) |
| | Diabetes (any type) | N (%) | 330 (1.3%) | 341 (1.3%) |
| | Hypothyroidism | N (%) | 1,111 (4.2%) | 1,154 (4.4%) |
| | Asthma | N (%) | 1,081 (4.1%) | 1,310 (5.0%) |
| | PCOS | N (%) | 927 (3.5%) | 879 (3.4%) |
| | Depressive disorder | N (%) | 2,415 (9.2%) | 2,488 (9.5%) |
| | HIV | N (%) | 5 (0.0%) | 5 (0.0%) |
| | Interstitial lung disease | N (%) | 31 (0.1%) | 40 (0.2%) |
| | Renal impairment | N (%) | 47 (0.2%) | 51 (0.2%) |
| | Pregnancy induced hypertension | N (%) | 658 (2.5%) | 613 (2.3%) |
| **Medications Prescribed (Last 180 Days)** | Antithrombotics | N (%) | 535 (2.0%) | 421 (1.6%) |
| | Immunosupressants | N (%) | 173 (0.7%) | 177 (0.7%) |
| | Progestogens | N (%) | 1,796 (6.9%) | 1,979 (7.6%) |
| | Estrogens | N (%) | 1,077 (4.1%) | 1,211 (4.6%) |
| | Antimycotics systemic | N (%) | 311 (1.2%) | 302 (1.2%) |
| | Antidepressants | N (%) | 2,304 (8.8%) | 2,491 (9.5%) |
| | Analgesics and antipyretics | N (%) | 3,675 (14.0%) | 3,767 (14.4%) |
| | Corticosteroids | N (%) | 439 (1.7%) | 434 (1.7%) |
| | Antihistamines systemic | N (%) | 5,799 (22.1%) | 6,038 (23.1%) |
| | Antibacterials systemic | N (%) | 2,766 (10.6%) | 2,629 (10.0%) |
| | Drugs ulcer and gords | N (%) | 1,249 (4.8%) | 1,099 (4.2%) |
| | Antiepileptics | N (%) | 217 (0.8%) | 225 (0.9%) |

| Covariate | Covariate level | Estimate | Cohort | |
|---|---|---|---|---|
| | | | Unexposed | Exposed |
| | Antiinflammatory antirehumatic | N (%) | 827 (3.2%) | 784 (3.0%) |

**Supplementary Table 7:** Baseline characteristics of the study population in the SIDIAP database, for the Primary (2-dose) Vaccination Schema vs. Unvaccinated analysis.

| Covariate | Covariate level | Estimate | Cohort | |
|---|---|---|---|---|
| | | | Unexposed | Exposed |
| **Number records** | - | N | 7,479 | 7,479 |
| **Age (Years)** | - | Median (Q25 - Q75) | 32.00 (28.00 - 36.00) | 32.00 (28.00 - 36.00) |
| **Age Group** | 12 to 24 | N (%) | 760 (10.2%) | 757 (10.1%) |
| | 25 to 39 | N (%) | 6,103 (81.6%) | 6,112 (81.7%) |
| | 40 to 55 | N (%) | 616 (8.2%) | 610 (8.2%) |
| **Gestational Trimester** | T1 | N (%) | 2,183 (29.2%) | 2,183 (29.2%) |
| | T2 | N (%) | 3,351 (44.8%) | 3,351 (44.8%) |
| | T3 | N (%) | 1,945 (26.0%) | 1,945 (26.0%) |
| **Vaccine Product** | Moderna | N (%) | - | 1,496 (20.0%) |
| | Pfizer | N (%) | - | 5,983 (80.0%) |
| **Previous Pregnancies** | - | Median (Q25 - Q75) | 2.00 (1.00 - 2.00) | 2.00 (2.00 - 2.00) |
| **Healthcare Visits (Past Year)** | - | Median (Q25 - Q75) | 20.00 (10.00 - 36.00) | 32.00 (16.00 - 52.00) |
| **Days of Prior Observation** | - | Median (Q25 - Q75) | 5,658 (2,816 - 5,700) | 5,664 (2,822 - 5,702) |
| **COVID-19 Infection (Any Time Prior)** | - | Median (Q25 - Q75) | 0.00 (0.00 - 0.00) | 0.00 (0.00 - 0.00) |
| **Other Vaccinations (Any Time Prior)** | Influenza | N (%) | 1,752 (23.4%) | 2,107 (28.2%) |
| | Tdap | N (%) | 3,366 (45.0%) | 3,329 (44.5%) |
| **Comorbidities (Any Time Prior)** | Asthma | N (%) | 370 (4.9%) | 455 (6.1%) |
| | Pneumonia | N (%) | 206 (2.8%) | 245 (3.3%) |

| Covariate | Covariate level | Estimate | Cohort | |
|---|---|---|---|---|
| | | | Unexposed | Exposed |
| | Interstitial lung disease | N (%) | 2 (0.0%) | 5 (0.1%) |
| | Stroke | N (%) | 6 (0.1%) | 8 (0.1%) |
| | PCOS | N (%) | 251 (3.4%) | 305 (4.1%) |
| | Chronic hypertension | N (%) | 155 (2.1%) | 178 (2.4%) |
| | Pregnancy induced hypertension | N (%) | 78 (1.0%) | 80 (1.1%) |
| | Anemia | N (%) | 1,876 (25.1%) | 1,647 (22.0%) |
| | Hypothyroidism | N (%) | 596 (8.0%) | 659 (8.8%) |
| | Depressive disorder | N (%) | 420 (5.6%) | 423 (5.7%) |
| | Heart conditions | N (%) | 142 (1.9%) | 176 (2.4%) |
| | Diabetes (any type) | N (%) | 402 (5.4%) | 442 (5.9%) |
| | HIV | N (%) | 4 (0.1%) | 8 (0.1%) |
| | Anxiety disorder | N (%) | 2,067 (27.6%) | 2,060 (27.5%) |
| | Renal impairment | N (%) | 9 (0.1%) | 22 (0.3%) |
| | Venous thromboembolism | N (%) | 49 (0.7%) | 48 (0.6%) |
| **Medications Prescribed (Last 180 Days)** | Estrogens | N (%) | 590 (7.9%) | 548 (7.3%) |
| | Antiinflammatory antirehumatic | N (%) | 1,186 (15.9%) | 1,215 (16.2%) |
| | Antithrombotics | N (%) | 347 (4.6%) | 283 (3.8%) |
| | Drugs ulcer and gords | N (%) | 439 (5.9%) | 444 (5.9%) |
| | Immunosupressants | N (%) | 9 (0.1%) | 13 (0.2%) |
| | Antidepressants | N (%) | 330 (4.4%) | 416 (5.6%) |
| | Progestogens | N (%) | 410 (5.5%) | 409 (5.5%) |
| | Antiepileptics | N (%) | 106 (1.4%) | 132 (1.8%) |
| | Analgesics and antipyretics | N (%) | 2,467 (33.0%) | 2,528 (33.8%) |

| Covariate | Covariate level | Estimate | Cohort | |
|---|---|---|---|---|
| | | | Unexposed | Exposed |
| | Antihistamines systemic | N (%) | 1,068 (14.3%) | 1,186 (15.9%) |
| | Antibacterials systemic | N (%) | 1,691 (22.6%) | 1,738 (23.2%) |
| | Corticosteroids | N (%) | 184 (2.5%) | 181 (2.4%) |
| | Antimycotics systemic | N (%) | 39 (0.5%) | 43 (0.6%) |

**Supplementary Table 8:** Baseline characteristics of the study population in the SIDIAP database, for the 1st 1st Booster (3rd dose) (3rd dose) vs. Primary (2-dose) Vaccination Schema analysis.

| Covariate | Covariate level | Estimate | Cohort | |
|---|---|---|---|---|
| | | | Unexposed | Exposed |
| **Number records** | - | N | 2,218 | 2,218 |
| **Age (Years)** | - | Median (Q25 - Q75) | 34.00 (31.00 - 37.00) | 34.00 (31.00 - 37.00) |
| **Age Group** | 12 to 24 | N (%) | 56 (2.5%) | 64 (2.9%) |
| | 25 to 39 | N (%) | 1,981 (89.3%) | 1,973 (89.0%) |
| | 40 to 55 | N (%) | 181 (8.2%) | 181 (8.2%) |
| **Gestational Trimester** | T1 | N (%) | 719 (32.4%) | 719 (32.4%) |
| | T2 | N (%) | 1,018 (45.9%) | 1,018 (45.9%) |
| | T3 | N (%) | 481 (21.7%) | 481 (21.7%) |
| **Vaccine Product** | Moderna | N (%) | - | 1,545 (69.7%) |
| | Pfizer | N (%) | - | 673 (30.3%) |
| **Previous Pregnancies** | - | Median (Q25 - Q75) | 2.00 (2.00 - 2.00) | 2.00 (2.00 - 2.00) |
| **Healthcare Visits (Past Year)** | - | Median (Q25 - Q75) | 26.00 (14.00 - 46.00) | 30.00 (16.00 - 52.00) |
| **Days of Prior Observation** | - | Median (Q25 - Q75) | 5,847 (4,332 - 5,876) | 5,842 (3,665 - 5,872) |
| **COVID-19 Infection (Any Time Prior)** | - | Median (Q25 - Q75) | 0.00 (0.00 - 0.00) | 0.00 (0.00 - 0.00) |
| **Other Vaccinations (Any Time Prior)** | Influenza | N (%) | 1,063 (47.9%) | 1,228 (55.4%) |
| | Tdap | N (%) | 934 (42.1%) | 887 (40.0%) |
| **Comorbidities (Any Time Prior)** | Asthma | N (%) | 133 (6.0%) | 145 (6.5%) |
| | Pneumonia | N (%) | 58 (2.6%) | 52 (2.3%) |
| | Interstitial lung disease | N (%) | 2 (0.1%) | 1 (0.0%) |
| | Stroke | N (%) | 1 (0.0%) | 0 (0.0%) |

| Covariate | Covariate level | Estimate | Cohort | |
|---|---|---|---|---|
| | | | Unexposed | Exposed |
| | PCOS | N (%) | 97 (4.4%) | 86 (3.9%) |
| | Chronic hypertension | N (%) | 35 (1.6%) | 53 (2.4%) |
| | Pregnancy induced hypertension | N (%) | 14 (0.6%) | 22 (1.0%) |
| | Anemia | N (%) | 443 (20.0%) | 413 (18.6%) |
| | Hypothyroidism | N (%) | 200 (9.0%) | 218 (9.8%) |
| | Depressive disorder | N (%) | 116 (5.2%) | 118 (5.3%) |
| | Heart conditions | N (%) | 55 (2.5%) | 49 (2.2%) |
| | Diabetes (any type) | N (%) | 89 (4.0%) | 111 (5.0%) |
| | HIV | N (%) | 0 (0.0%) | 1 (0.0%) |
| | Anxiety disorder | N (%) | 609 (27.5%) | 551 (24.8%) |
| | Renal impairment | N (%) | 6 (0.3%) | 5 (0.2%) |
| | Venous thromboembolism | N (%) | 10 (0.5%) | 16 (0.7%) |
| **Medications Prescribed (Last 180 Days)** | Estrogens | N (%) | 128 (5.8%) | 130 (5.9%) |
| | Antiinflammatory antirehumatic | N (%) | 368 (16.6%) | 372 (16.8%) |
| | Antithrombotics | N (%) | 164 (7.4%) | 122 (5.5%) |
| | Drugs ulcer and gords | N (%) | 135 (6.1%) | 117 (5.3%) |
| | Immunosupressants | N (%) | 1 (0.0%) | 1 (0.0%) |
| | Antidepressants | N (%) | 112 (5.0%) | 106 (4.8%) |
| | Progestogens | N (%) | 126 (5.7%) | 126 (5.7%) |
| | Antiepileptics | N (%) | 28 (1.3%) | 35 (1.6%) |
| | Analgesics and antipyretics | N (%) | 720 (32.5%) | 705 (31.8%) |
| | Antihistamines systemic | N (%) | 316 (14.2%) | 366 (16.5%) |
| | Antibacterials systemic | N (%) | 463 (20.9%) | 410 (18.5%) |
| | Corticosteroids | N (%) | 61 (2.8%) | 68 (3.1%) |
| | Antimycotics systemic | N (%) | 16 (0.7%) | 10 (0.5%) |

**Supplementary Table 9:** Baseline characteristics of the study population in the UiO database, for the Primary (2-dose) Vaccination Schema vs. Unvaccinated analysis.

| Covariate | Covariate level | Estimate | Cohort | |
|---|---|---|---|---|
| | | | Unexposed | Exposed |
| **Number records** | - | N | 15,443 | 15,443 |
| **Age (Years)** | - | Median (Q25 - Q75) | 31.00 (28.00 - 34.00) | 31.00 (28.00 - 34.00) |
| **Age Group** | 12 to 24 | N (%) | 1,360 (8.8%) | 1,367 (8.9%) |
| | 25 to 39 | N (%) | 13,512 (87.5%) | 13,507 (87.5%) |
| | 40 to 55 | N (%) | 571 (3.7%) | 569 (3.7%) |
| **Gestational Trimester** | T1 | N (%) | 5,201 (33.7%) | 5,201 (33.7%) |
| | T2 | N (%) | 6,421 (41.6%) | 6,421 (41.6%) |
| | T3 | N (%) | 3,821 (24.7%) | 3,821 (24.7%) |
| **Vaccine Product** | Moderna | N (%) | - | 5,248 (34.0%) |
| | Pfizer | N (%) | - | 10,195 (66.0%) |
| **Previous Pregnancies** | - | Median (Q25 - Q75) | 2.00 (1.00 - 2.00) | 2.00 (2.00 - 2.00) |
| **Healthcare Visits (Past Year)** | - | Median (Q25 - Q75) | 20.00 (11.00 - 34.00) | 30.00 (16.00 - 48.00) |
| **Days of Prior Observation** | - | Median (Q25 - Q75) | 4,964 (4,859 - 4,992) | 4,979 (4,923 - 4,993) |
| **COVID-19 Infection (Any Time Prior)** | - | Median (Q25 - Q75) | 0.00 (0.00 - 0.00) | 0.00 (0.00 - 0.00) |
| **Other Vaccinations (Any Time Prior)** | Influenza | N (%) | 3,154 (20.4%) | 4,307 (27.9%) |
| | Tdap | N (%) | 97 (0.6%) | 90 (0.6%) |
| **Comorbidities (Any Time Prior)** | Diabetes (any type) | N (%) | 822 (5.3%) | 821 (5.3%) |
| | Chronic hypertension | N (%) | 1,049 (6.8%) | 1,122 (7.3%) |

| Covariate | Covariate level | Estimate | Cohort | |
|---|---|---|---|---|
| | | | Unexposed | Exposed |
| | Anemia | N (%) | 2,892 (18.7%) | 2,509 (16.2%) |
| | Anxiety disorder | N (%) | 1,745 (11.3%) | 1,722 (11.2%) |
| | Venous thromboembolism | N (%) | 243 (1.6%) | 250 (1.6%) |
| | Stroke | N (%) | 38 (0.2%) | 37 (0.2%) |
| | Renal impairment | N (%) | 80 (0.5%) | 63 (0.4%) |
| | Heart conditions | N (%) | 796 (5.2%) | 839 (5.4%) |
| | Asthma | N (%) | 2,004 (13.0%) | 2,207 (14.3%) |
| | Hypothyroidism | N (%) | 890 (5.8%) | 865 (5.6%) |
| | Pneumonia | N (%) | 1,203 (7.8%) | 1,213 (7.9%) |
| | HIV | N (%) | 19 (0.1%) | 15 (0.1%) |
| | Pregnancy induced hypertension | N (%) | 850 (5.5%) | 901 (5.8%) |
| | PCOS | N (%) | 534 (3.5%) | 618 (4.0%) |
| | Interstitial lung disease | N (%) | 4 (0.0%) | 6 (0.0%) |
| | Depressive disorder | N (%) | 1,714 (11.1%) | 1,757 (11.4%) |
| **Medications Prescribed (Last 180 Days)** | Antihistamines systemic | N (%) | 2,768 (17.9%) | 3,239 (21.0%) |
| | Antibacterials systemic | N (%) | 2,095 (13.6%) | 1,948 (12.6%) |
| | Antithrombotics | N (%) | 718 (4.6%) | 774 (5.0%) |
| | Antidepressants | N (%) | 622 (4.0%) | 718 (4.6%) |
| | Antiinflammatory antirehumatic | N (%) | 1,150 (7.4%) | 1,203 (7.8%) |
| | Estrogens | N (%) | 1,835 (11.9%) | 2,063 (13.4%) |
| | Drugs ulcer and gords | N (%) | 693 (4.5%) | 676 (4.4%) |
| | Antiepileptics | N (%) | 146 (0.9%) | 166 (1.1%) |

| Covariate | Covariate level | Estimate | Cohort | |
|---|---|---|---|---|
| | | | Unexposed | Exposed |
| | Immunosupressants | N (%) | 74 (0.5%) | 114 (0.7%) |
| | Antimycotics systemic | N (%) | 134 (0.9%) | 146 (0.9%) |
| | Progestogens | N (%) | 1,512 (9.8%) | 1,536 (9.9%) |
| | Corticosteroids | N (%) | 292 (1.9%) | 308 (2.0%) |
| | Analgesics and antipyretics | N (%) | 1,173 (7.6%) | 1,243 (8.0%) |

**Supplementary Table 10:** Baseline characteristics of the study population in the UiO database, for the 1st 1st Booster (3rd dose) (3rd dose) vs. Primary (2-dose) Vaccination Schema analysis.

| Covariate | Covariate level | Estimate | Cohort | |
|---|---|---|---|---|
| | | | Unexposed | Exposed |
| **Number records** | - | N | 10,791 | 10,791 |
| **Age (Years)** | - | Median (Q25 - Q75) | 31.00 (28.00 - 34.00) | 31.00 (28.00 - 34.00) |
| **Age Group** | 12 to 24 | N (%) | 727 (6.7%) | 709 (6.6%) |
| | 25 to 39 | N (%) | 9,770 (90.5%) | 9,778 (90.6%) |
| | 40 to 55 | N (%) | 294 (2.7%) | 304 (2.8%) |
| **Gestational Trimester** | T1 | N (%) | 3,372 (31.2%) | 3,372 (31.2%) |
| | T2 | N (%) | 6,200 (57.5%) | 6,200 (57.5%) |
| | T3 | N (%) | 1,219 (11.3%) | 1,219 (11.3%) |
| **Vaccine Product** | Moderna | N (%) | - | 2,020 (18.7%) |
| | Pfizer | N (%) | - | 8,771 (81.3%) |
| **Previous Pregnancies** | - | Median (Q25 - Q75) | 2.00 (2.00 - 2.00) | 2.00 (2.00 - 2.00) |
| **Healthcare Visits (Past Year)** | - | Median (Q25 - Q75) | 24.00 (12.00 - 40.00) | 26.00 (16.00 - 44.00) |
| **Days of Prior Observation** | - | Median (Q25 - Q75) | 5,126 (5,092 - 5,144) | 5,127 (5,098 - 5,146) |
| **COVID-19 Infection (Any Time Prior)** | - | Median (Q25 - Q75) | 0.00 (0.00 - 0.00) | 0.00 (0.00 - 0.00) |
| **Other Vaccinations (Any Time Prior)** | Influenza | N (%) | 5,481 (50.8%) | 6,534 (60.6%) |
| | Tdap | N (%) | 84 (0.8%) | 69 (0.6%) |
| **Comorbidities (Any Time Prior)** | Diabetes (any type) | N (%) | 534 (4.9%) | 518 (4.8%) |
| | Chronic hypertension | N (%) | 806 (7.5%) | 816 (7.6%) |

| Covariate | Covariate level | Estimate | Cohort | |
|---|---|---|---|---|
| | | | Unexposed | Exposed |
| | Anemia | N (%) | 1,782 (16.5%) | 1,690 (15.7%) |
| | Anxiety disorder | N (%) | 1,083 (10.0%) | 1,085 (10.1%) |
| | Venous thromboembolism | N (%) | 185 (1.7%) | 176 (1.6%) |
| | Stroke | N (%) | 35 (0.3%) | 36 (0.3%) |
| | Renal impairment | N (%) | 58 (0.5%) | 56 (0.5%) |
| | Heart conditions | N (%) | 654 (6.1%) | 623 (5.8%) |
| | Asthma | N (%) | 1,589 (14.7%) | 1,700 (15.8%) |
| | Hypothyroidism | N (%) | 625 (5.8%) | 670 (6.2%) |
| | Pneumonia | N (%) | 860 (8.0%) | 869 (8.1%) |
| | HIV | N (%) | 3 (0.0%) | 5 (0.0%) |
| | Pregnancy induced hypertension | N (%) | 642 (5.9%) | 647 (6.0%) |
| | PCOS | N (%) | 426 (3.9%) | 448 (4.2%) |
| | Interstitial lung disease | N (%) | 5 (0.0%) | 3 (0.0%) |
| | Depressive disorder | N (%) | 1,138 (10.5%) | 1,186 (11.0%) |
| **Medications Prescribed (Last 180 Days)** | Antihistamines systemic | N (%) | 2,291 (21.2%) | 2,408 (22.3%) |
| | Antibacterials systemic | N (%) | 1,652 (15.3%) | 1,614 (15.0%) |
| | Antithrombotics | N (%) | 511 (4.7%) | 538 (5.0%) |
| | Antidepressants | N (%) | 413 (3.8%) | 424 (3.9%) |
| | Antiinflammatory antirehumatic | N (%) | 808 (7.5%) | 782 (7.2%) |
| | Estrogens | N (%) | 1,443 (13.4%) | 1,623 (15.0%) |
| | Drugs ulcer and gords | N (%) | 468 (4.3%) | 443 (4.1%) |
| | Antiepileptics | N (%) | 111 (1.0%) | 116 (1.1%) |

| Covariate | Covariate level | Estimate | Cohort | |
|---|---|---|---|---|
| | | | Unexposed | Exposed |
| | Immunosupressants | N (%) | 60 (0.6%) | 91 (0.8%) |
| | Antimycotics systemic | N (%) | 102 (0.9%) | 99 (0.9%) |
| | Progestogens | N (%) | 1,004 (9.3%) | 975 (9.0%) |
| | Corticosteroids | N (%) | 199 (1.8%) | 210 (1.9%) |
| | Analgesics and antipyretics | N (%) | 910 (8.4%) | 914 (8.5%) |

**Supplementary Table 11:** Additional information on follow-up and reasons for censoring. CPRD GOLD database, for the Primary (2-dose) Vaccination Schema vs. Unvaccinated analysis.

| Cohort | Reason | N | Days, Median (Q25-Q75) |
|---|---|---|---|
| Exposed | End of pregnancy | 229 (4.177%) | 42.00 (19.00 - 98.00) |
| | Next COVID-19 vaccine dose | 8 (0.146%) | 146.50 (142.25 - 178.50) |
| | Pair matched next COVID-19 vaccine dose | 357 (6.512%) | 20.00 (11.00 - 31.00) |
| | No second dose | 226 (4.123%) | 42.00 (42.00 - 42.00) |
| | Second dose before recommended time | <5 | - |
| | Second dose after recommended time | 1,913 (34.896%) | 42.00 (42.00 - 42.00) |
| | Second dose after recommended time; unexposed 1st dose | <5 | - |
| Unexposed | End of pregnancy | 213 (3.885%) | 43.00 (30.00 - 97.00) |
| | Next COVID-19 vaccine dose | 344 (6.275%) | 19.00 (10.00 - 31.00) |
| | Pair matched next COVID-19 vaccine dose | 12 (0.219%) | 164.50 (143.75 - 185.50) |
| | No second dose | 227 (4.141%) | 42.00 (42.00 - 42.00) |
| | Second dose before recommended time | <5 | - |
| | Second dose after recommended time | 1,937 (35.334%) | 42.00 (42.00 - 42.00) |
| | Second dose after recommended time; unexposed 1st dose | <5 | - |

**Supplementary Table 12:** Additional information on follow-up and reasons for censoring. CPRD GOLD database, for the 1st Booster (3rd dose) vs. Primary (2-dose) Vaccination Schema analysis.

| Cohort | Reason | N | Days, Median (Q25-Q75) |
|---|---|---|---|
| Exposed | End of pregnancy | 1,030 (27.22%) | 163.00 (103.00 - 221.00) |
| | Next COVID-19 vaccine dose | 6 (0.159%) | 119.50 (44.75 - 135.75) |
| | Pair matched next COVID-19 vaccine dose | 856 (22.622%) | 17.00 (8.00 - 37.00) |
| Unexposed | End of pregnancy | 1,039 (27.458%) | 162.00 (101.50 - 220.00) |
| | Next COVID-19 vaccine dose | 847 (22.384%) | 17.00 (7.00 - 36.00) |
| | Pair matched next COVID-19 vaccine dose | 6 (0.159%) | 119.50 (44.75 - 135.75) |

**Supplementary Table 13:** Additional information on follow-up and reasons for censoring. SCIFI-PEARL database, for the Primary (2-dose) Vaccination Schema vs. Unvaccinated analysis.

| Cohort | Reason | N | Days, Median (Q25-Q75) |
|---|---|---|---|
| **Exposed** | End of pregnancy | 11,568 (15.891%) | 123.00 (77.00 - 166.00) |
| | Next COVID-19 vaccine dose | 755 (1.037%) | 161.00 (147.50 - 179.00) |
| | Pair matched next COVID-19 vaccine dose | 10,084 (13.852%) | 21.00 (10.00 - 41.00) |
| | No second dose | 1,270 (1.745%) | 42.00 (42.00 - 42.00) |
| | Second dose before recommended time | 11 (0.015%) | 13.00 (9.00 - 17.50) |
| | Second dose after recommended time | 12,655 (17.384%) | 42.00 (42.00 - 42.00) |
| | Second dose after recommended time; unexposed 1st dose | 29 (0.04%) | 42.00 (42.00 - 42.00) |
| | Unexposed 1st dose; no second dose | <5 | - |
| | Unexposed 1st dose; second dose after recommended time | 23 (0.032%) | 42.00 (42.00 - 42.00) |
| **Unexposed** | End of pregnancy | 11,778 (16.179%) | 123.00 (77.00 - 166.00) |
| | Next COVID-19 vaccine dose | 9,773 (13.425%) | 20.00 (10.00 - 39.00) |
| | Pair matched next COVID-19 vaccine dose | 765 (1.051%) | 161.00 (148.00 - 180.00) |
| | No second dose | 1,282 (1.761%) | 42.00 (42.00 - 42.00) |
| | Second dose before recommended time | 11 (0.015%) | 13.00 (9.00 - 17.50) |
| | Second dose after recommended time | 12,739 (17.5%) | 42.00 (42.00 - 42.00) |
| | Second dose after recommended time; unexposed 1st dose | 27 (0.037%) | 42.00 (42.00 - 42.00) |
| | Unexposed 1st dose; no second dose | <5 | - |

| Cohort | Reason | N | Days, Median (Q25-Q75) |
|---|---|---|---|
| | Unexposed 1st dose; second dose after recommended time | 20 (0.027%) | 42.00 (42.00 - 42.00) |

**Supplementary Table 14:** Additional information on follow-up and reasons for censoring. SCIFI-PEARL database, for the 1st Booster (3rd dose) vs. Primary (2-dose) Vaccination Schema analysis.

| Cohort | Reason | N | Days, Median (Q25-Q75) |
|---|---|---|---|
| **Exposed** | End of pregnancy | 15,707 (29.981%) | 138.00 (98.00 - 174.00) |
| | Next COVID-19 vaccine dose | 556 (1.061%) | 168.00 (138.00 - 218.00) |
| | Pair matched next COVID-19 vaccine dose | 9,932 (18.958%) | 21.00 (10.00 - 48.00) |
| **Unexposed** | End of pregnancy | 15,764 (30.09%) | 138.00 (97.00 - 174.00) |
| | Next COVID-19 vaccine dose | 9,901 (18.899%) | 21.00 (9.00 - 48.00) |
| | Pair matched next COVID-19 vaccine dose | 530 (1.012%) | 169.00 (137.25 - 218.00) |

**Supplementary Table 15:** Additional information on follow-up and reasons for censoring. SIDIAP database, for the Primary (2-dose) Vaccination Schema vs. Unvaccinated analysis.

| Cohort | Reason | N | Days, Median (Q25-Q75) |
|---|---|---|---|
| Exposed | End of pregnancy | 3,549 (23.726%) | 113.00 (70.00 - 162.00) |
| | Next COVID-19 vaccine dose | 81 (0.542%) | 199.00 (175.00 - 209.00) |
| | Pair matched next COVID-19 vaccine dose | 2,612 (17.462%) | 27.00 (12.00 - 55.00) |
| | No second dose | 711 (4.753%) | 42.00 (42.00 - 49.00) |
| | Second dose before recommended time | <5 | - |
| | Second dose after recommended time | 520 (3.476%) | 42.00 (42.00 - 42.00) |
| | Second dose after recommended time; unexposed 1st dose | <5 | - |
| | Unexposed 1st dose; no second dose | <5 | - |
| Unexposed | End of pregnancy | 3,625 (24.235%) | 113.00 (70.00 - 162.00) |
| | Next COVID-19 vaccine dose | 2,509 (16.774%) | 25.00 (12.00 - 51.00) |
| | Pair matched next COVID-19 vaccine dose | 90 (0.602%) | 199.50 (175.25 - 209.75) |
| | No second dose | 717 (4.793%) | 42.00 (42.00 - 49.00) |
| | Second dose before recommended time | 5 (0.033%) | 15.00 (14.00 - 15.00) |
| | Second dose after recommended time | 531 (3.55%) | 42.00 (42.00 - 42.00) |
| | Second dose after recommended time; unexposed 1st dose | <5 | - |
| | Unexposed 1st dose; no second dose | <5 | - |

**Supplementary Table 16:** Additional information on follow-up and reasons for censoring. SIDIAP database, for the 1st Booster (3rd dose) vs. Primary (2-dose) Vaccination Schema analysis.

| Cohort | Reason | N | Days, Median (Q25-Q75) |
|---|---|---|---|
| Exposed | End of pregnancy | 1,736 (39.134%) | 139.50 (90.00 - 192.00) |
| | Next COVID-19 vaccine dose | 13 (0.293%) | 182.00 (154.00 - 195.00) |
| | Pair matched next COVID-19 vaccine dose | 469 (10.573%) | 19.00 (8.00 - 42.00) |
| Unexposed | End of pregnancy | 1,753 (39.518%) | 139.00 (90.00 - 196.00) |
| | Next COVID-19 vaccine dose | 453 (10.212%) | 18.00 (8.00 - 40.00) |
| | Pair matched next COVID-19 vaccine dose | 12 (0.271%) | 171.50 (152.50 - 196.75) |

**Supplementary Table 17:** Additional information on follow-up and reasons for censoring. UiO database, for the Primary (2-dose) Vaccination Schema vs. Unvaccinated analysis.

| Cohort | Reason | N | Days, Median (Q25-Q75) |
|---|---|---|---|
| **Exposed** | End of pregnancy | 5,728 (18.546%) | 75.00 (42.00 - 125.00) |
| | Next COVID-19 vaccine dose | 115 (0.372%) | 171.00 (167.00 - 178.50) |
| | Pair matched next COVID-19 vaccine dose | 5,123 (16.587%) | 22.00 (9.00 - 60.00) |
| | No second dose | 669 (2.166%) | 42.00 (42.00 - 42.00) |
| | Second dose before recommended time | 15 (0.049%) | 20.00 (20.00 - 21.00) |
| | Second dose after recommended time | 3,777 (12.229%) | 42.00 (42.00 - 42.00) |
| | Second dose after recommended time; unexposed 1st dose | 16 (0.052%) | 42.00 (42.00 - 42.00) |
| **Unexposed** | End of pregnancy | 5,259 (17.027%) | 87.00 (55.50 - 136.00) |
| | Next COVID-19 vaccine dose | 5,130 (16.609%) | 22.00 (9.00 - 62.00) |
| | Pair matched next COVID-19 vaccine dose | 210 (0.68%) | 175.50 (168.00 - 188.00) |
| | No second dose | 718 (2.325%) | 42.00 (42.00 - 42.00) |
| | Second dose before recommended time | 15 (0.049%) | 20.00 (20.00 - 21.00) |
| | Second dose after recommended time | 4,099 (13.271%) | 42.00 (42.00 - 42.00) |
| | Second dose after recommended time; unexposed 1st dose | 11 (0.036%) | 42.00 (42.00 - 42.00) |
| | Exposed 3rd dose; unexposed 1st dose | <5 | - |

**Supplementary Table 18:** Additional information on follow-up and reasons for censoring. UiO database, for the 1st Booster (3rd dose) vs. Primary (2-dose) Vaccination Schema analysis.

| Cohort | Reason | N | Days, Median (Q25-Q75) |
|---|---|---|---|
| **Exposed** | End of pregnancy | 6,956 (32.231%) | 149.00 (97.00 - 184.00) |
| | Next COVID-19 vaccine dose | 22 (0.102%) | 168.00 (140.25 - 240.50) |
| | Pair matched next COVID-19 vaccine dose | 3,813 (17.668%) | 14.00 (7.00 - 27.00) |
| **Unexposed** | End of pregnancy | 6,918 (32.054%) | 154.00 (109.00 - 190.00) |
| | Next COVID-19 vaccine dose | 3,855 (17.862%) | 14.00 (7.00 - 27.00) |
| | Pair matched next COVID-19 vaccine dose | 18 (0.083%) | 158.50 (124.00 - 178.50) |

**Supplementary Figure 5:** Distribution of index dates (enrollment date) across all databases and analyses.

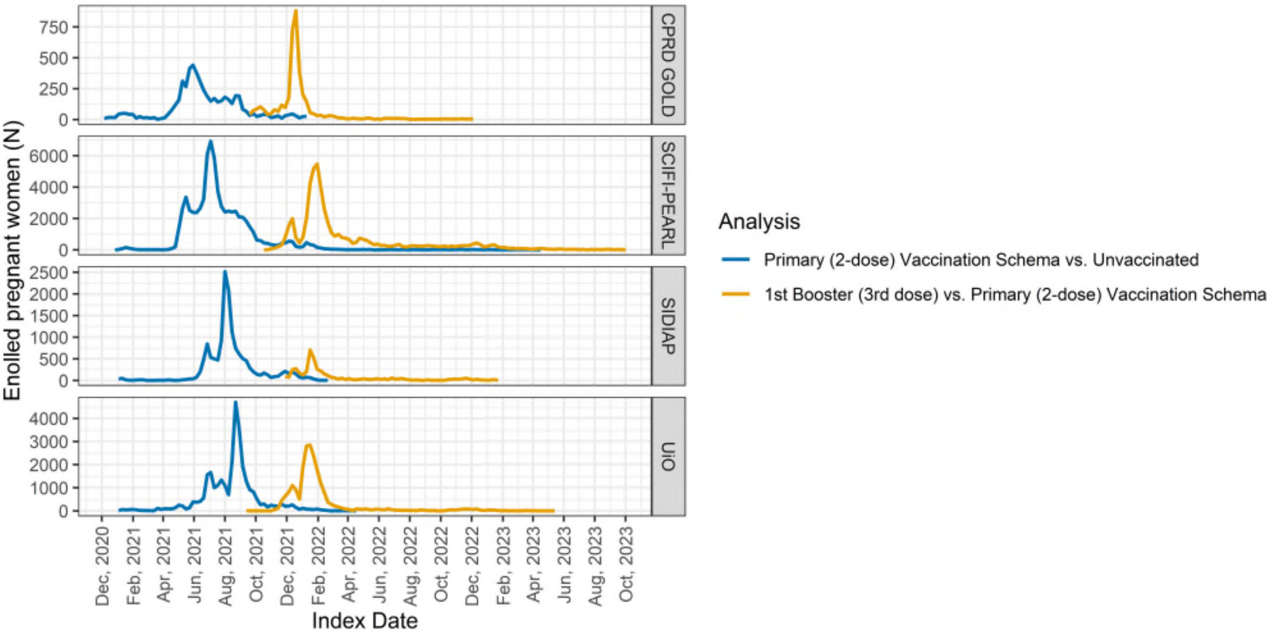

**Supplementary Figure 6:** Hazard Ratios for COVID-19 Infection and COVID-19 related Hospitalisation (Primary (2-dose) Vaccination Schema vs. Unvaccinated, and 1st Booster (3rd dose) vs. Primary (2-dose) Vaccination Schema) stratified by vaccine brand. Estimates across databases were pooled using random-effects meta-analysis.

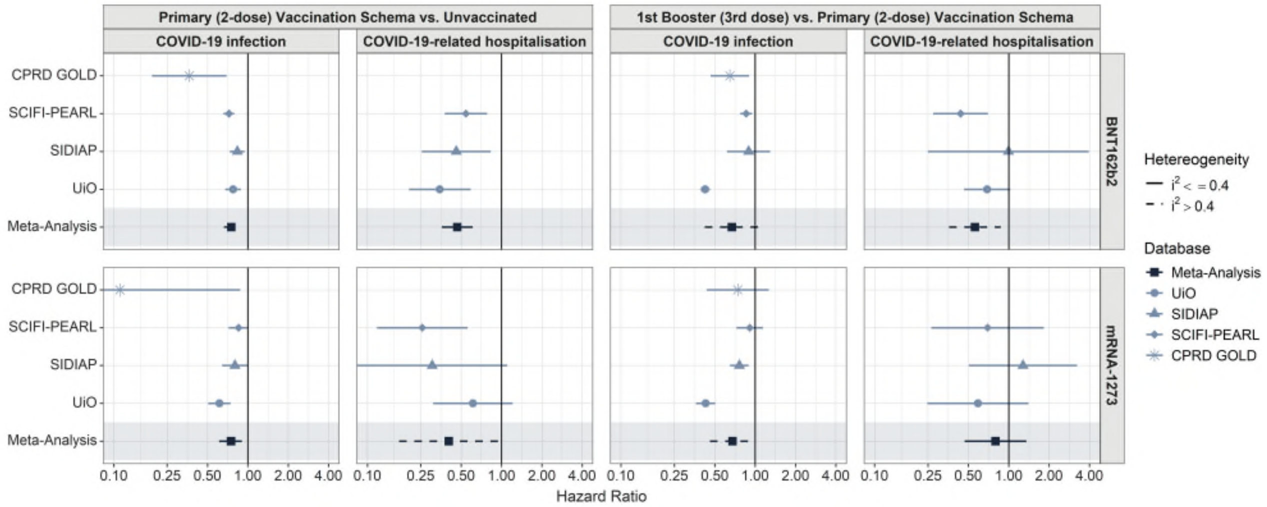

**Supplementary Figure 7:** Kaplan-Meier curves for COVID-19 infection, across databases, and analysis

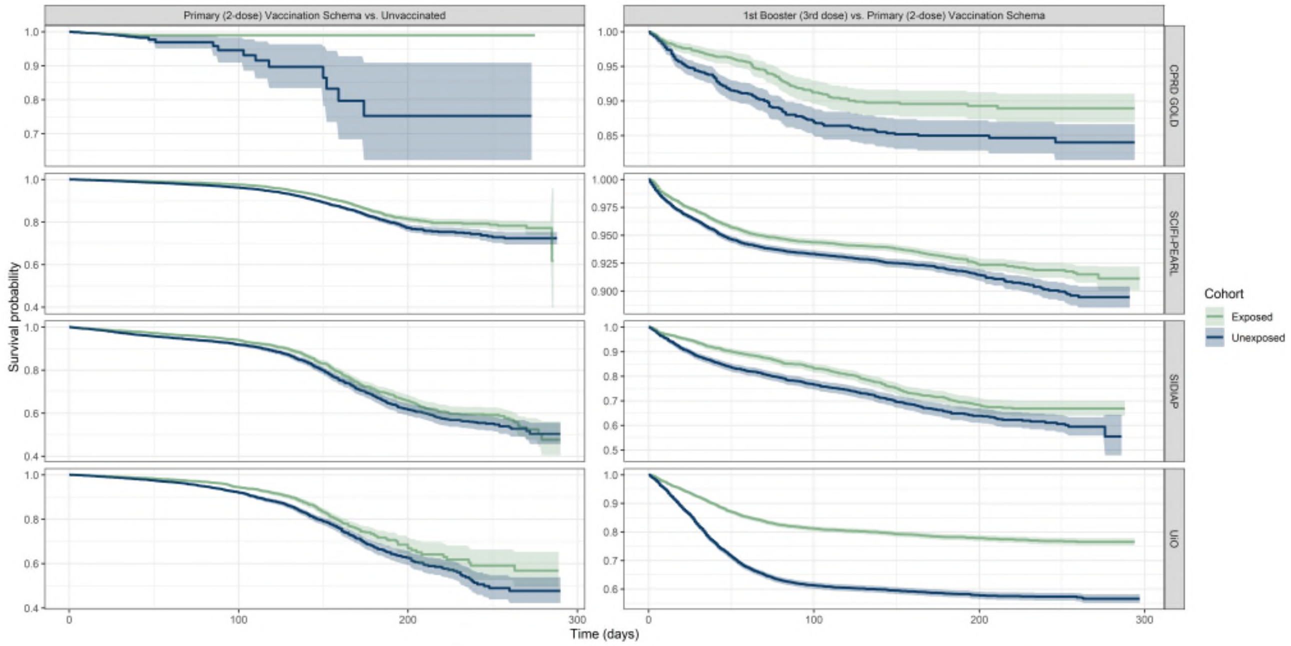

**Supplementary Figure 8:** Kaplan-Meier curves for COVID-19-related hospitalisation, across databases, and analysis

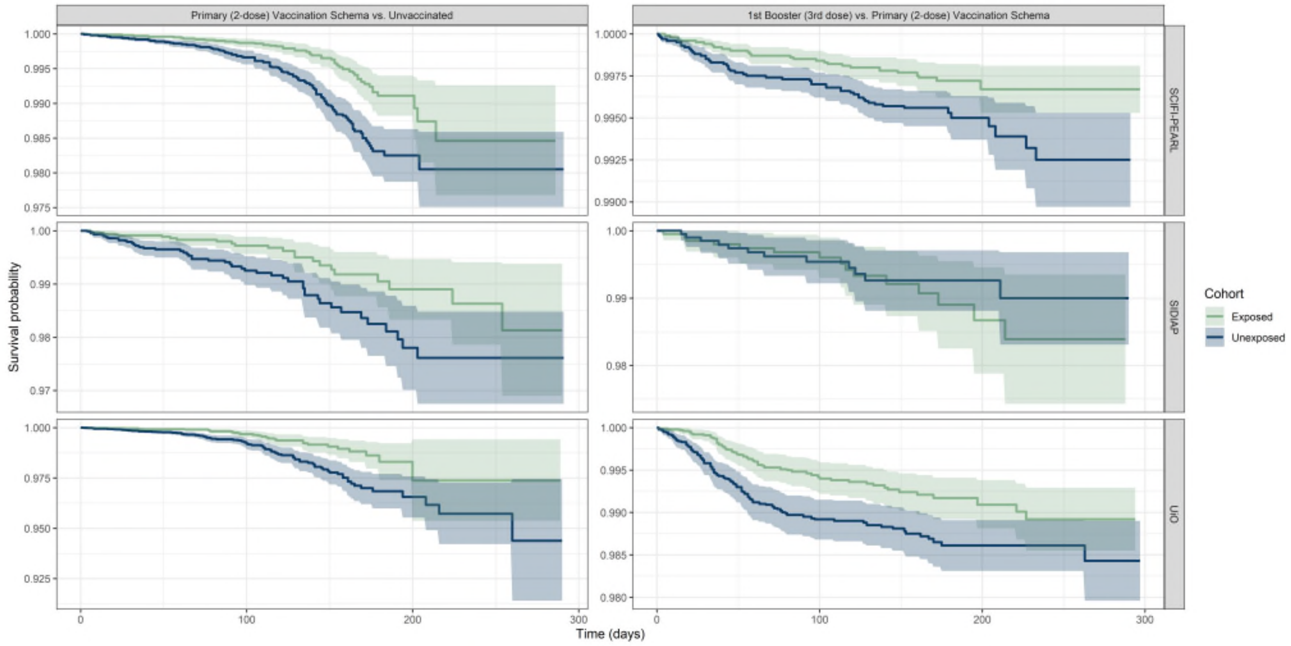

**Supplementary Figure 9:** Log-Log plots for COVID-19 infection, across databases, and analysis

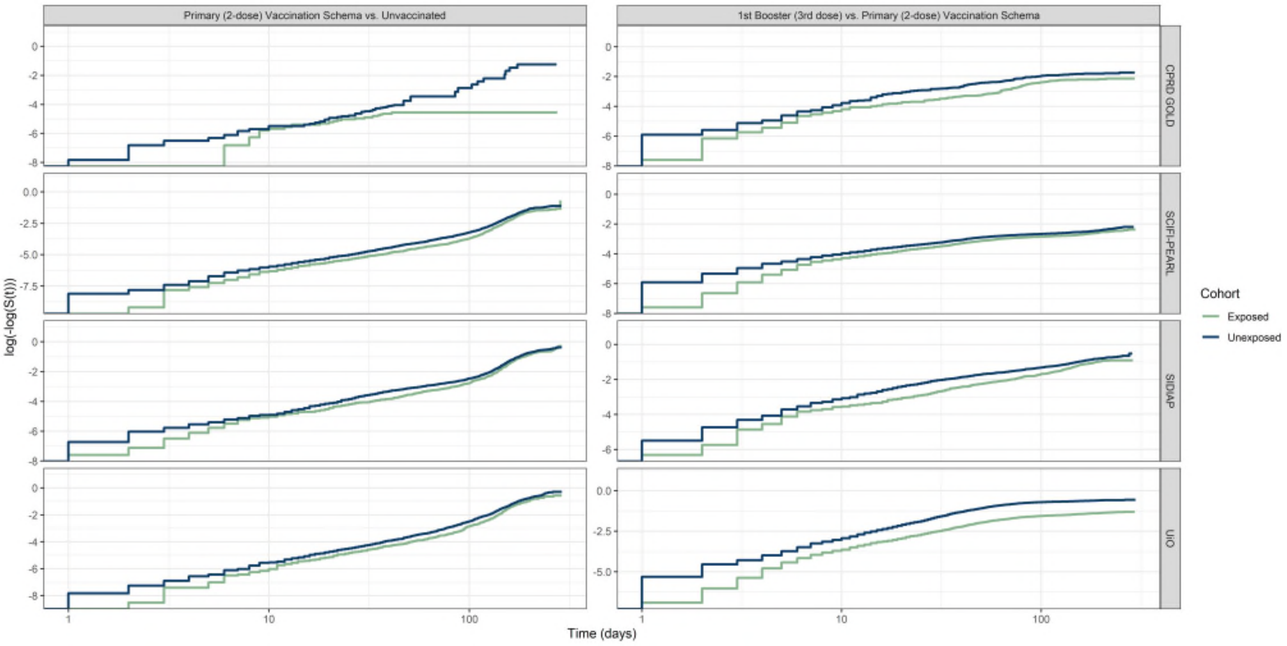

**Supplementary Figure 10:** Log-Log plots for COVID-19-related hospitalisation, across databases, and analysis

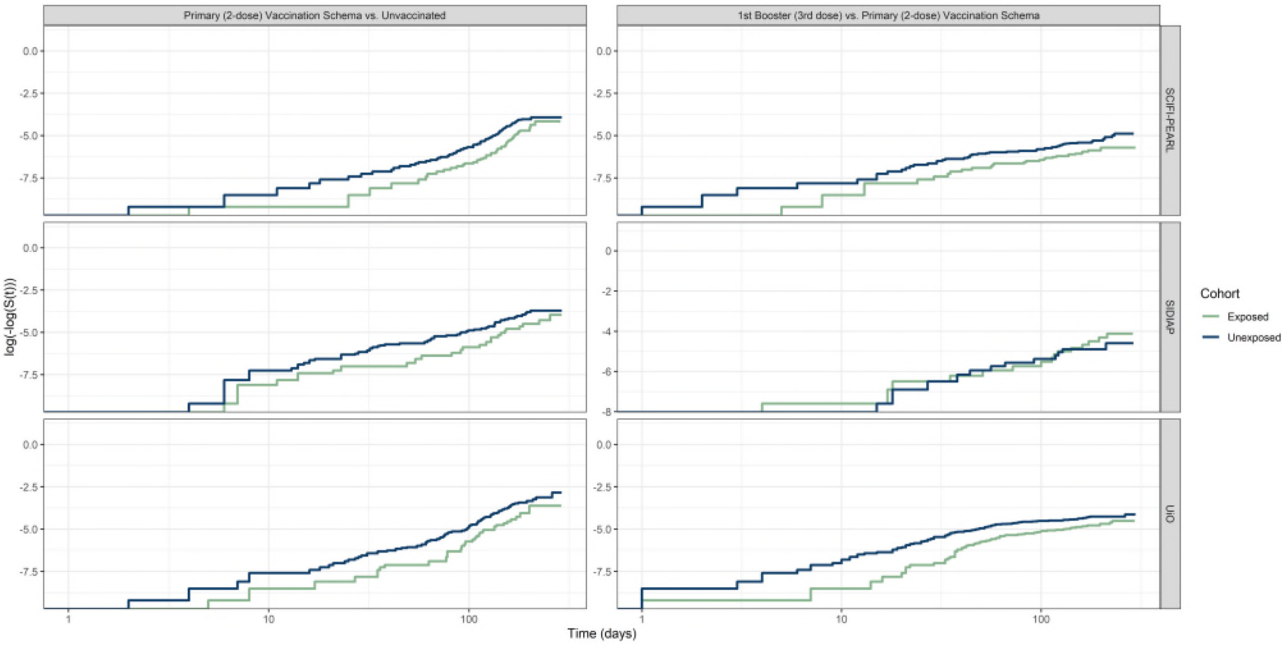

**Supplementary Figure 11:** Meta-Analysed Hazard Ratio estimates against COVID-19 infection and COVID-19 related hospitalisation across different time-windows post-vaccination.

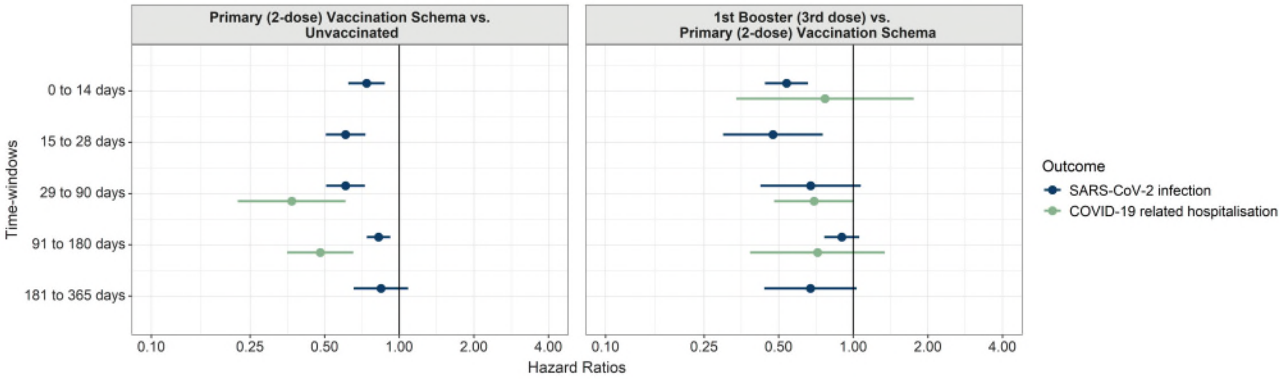

**Supplementary Figure 12:** Negative Control Outcomes results for SCIFI-PEARL database.

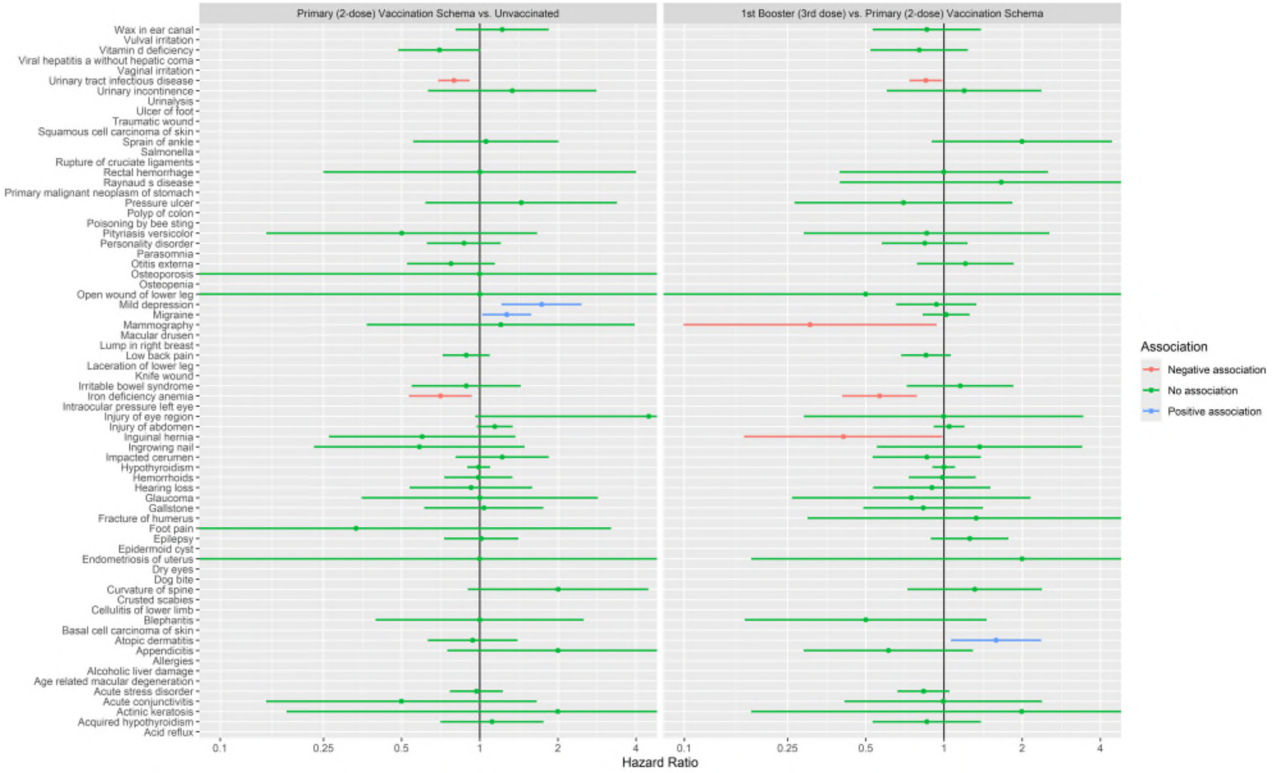

**Supplementary Figure 13:** Negative Control Outcomes results for UiO database.

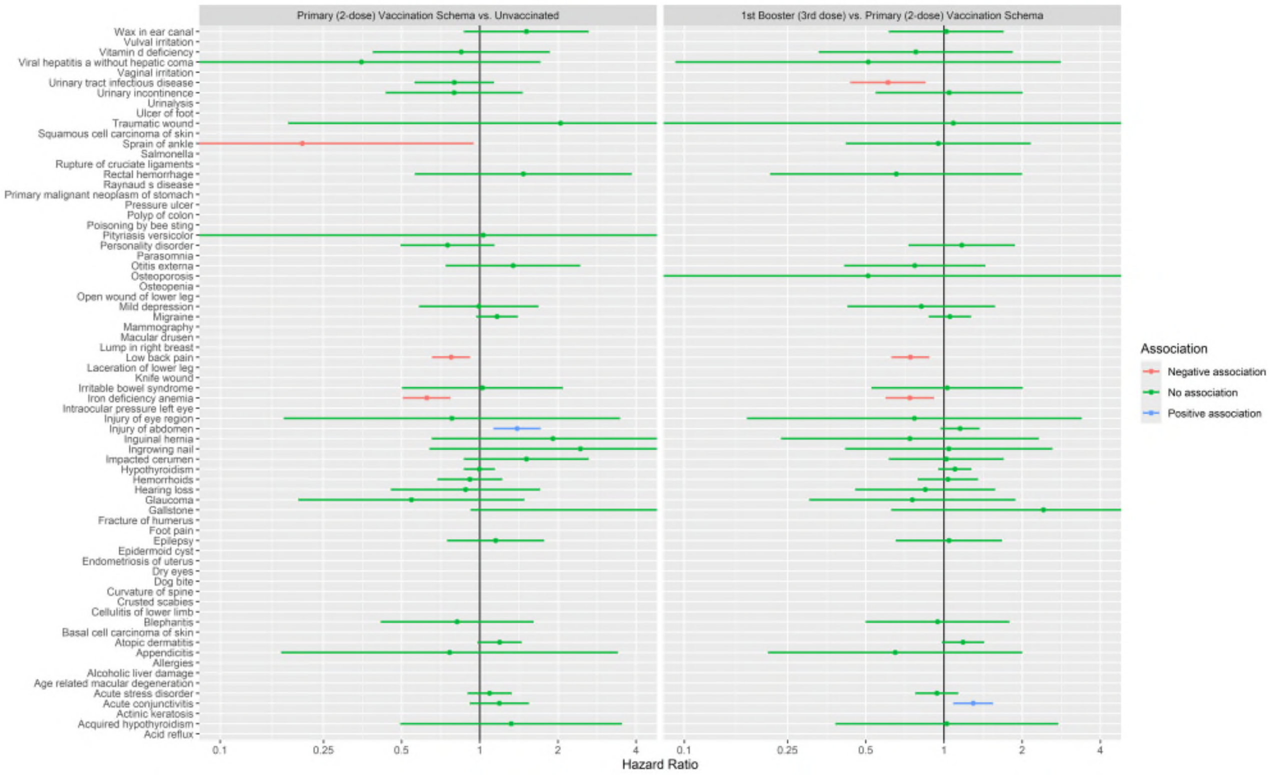

**Supplementary Figure 14:** Negative Control Outcomes results for SIDIAP database.

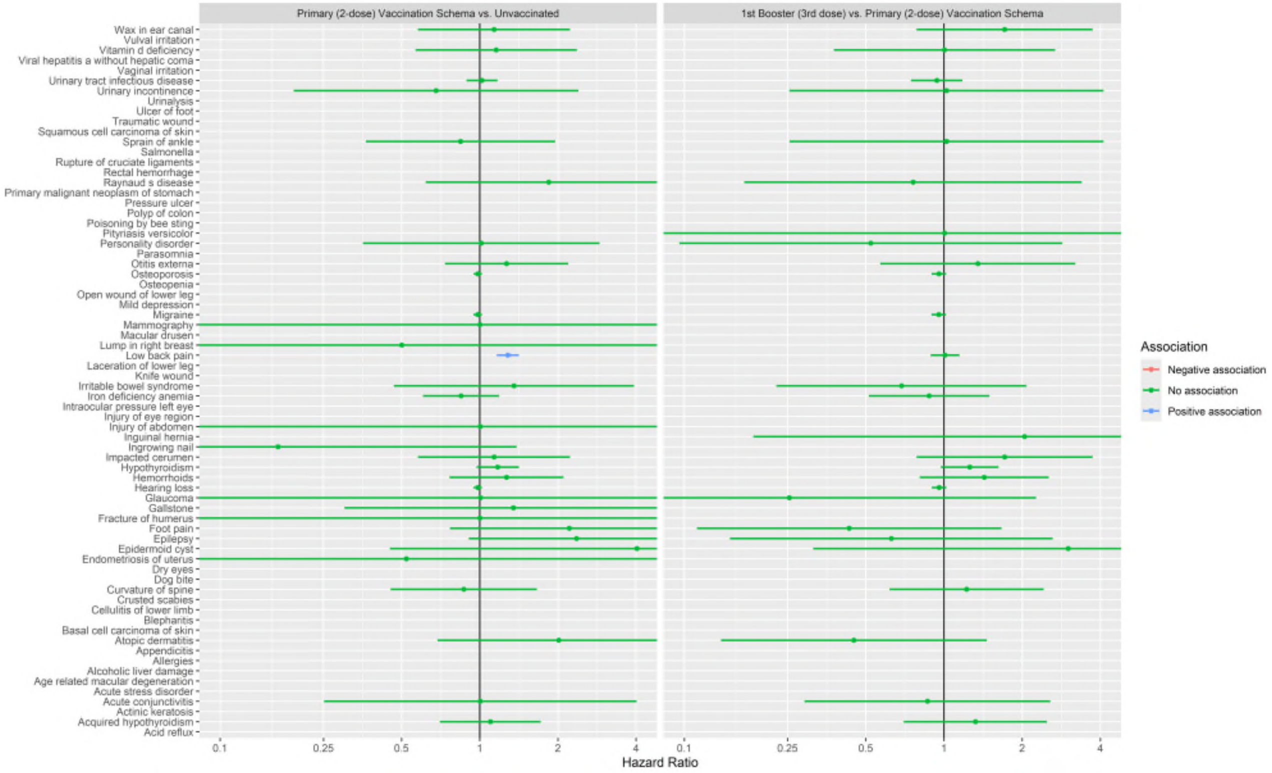

**Supplementary Figure 15:** Negative Control Outcomes results for CPRD GOLD database.

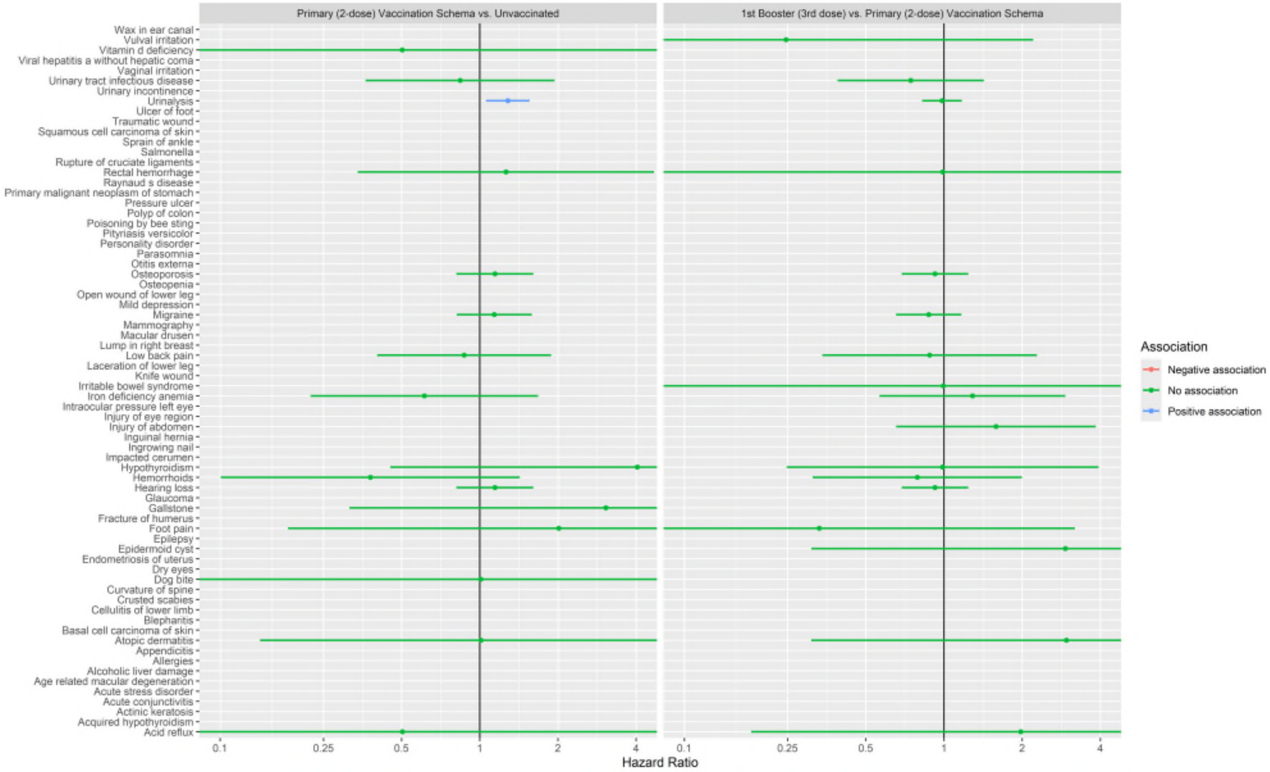

**Supplementary Figure 16:** Vaccine effectiveness estimates for Primary (2-dose) Vaccination Schema vs. Unvaccinated, when follow-up is not stopped at pregnancy end date.

| Database | Exposed | | Unexposed | | HR (95% CI) | VE | i2 |
|---|---|---|---|---|---|---|---|
| | Population (N) | Events (N(%)) | Population (N) | Events (N(%)) | | | |
| **COVID-19 infection** | | | | | | | |
| CPRD GOLD | 2,581 | 17 (0.66%) | 2,581 | 51 (1.98%) | 0.32 (0.19 - 0.56) | 67.86% | |
| SCIFI-PEARL | 32,429 | 1,382 (4.26%) | 32,429 | 1,737 (5.36%) | 0.78 (0.73 - 0.84) | 22.15% | |
| SIDIAP | 6,530 | 970 (14.85%) | 6,530 | 1,060 (16.23%) | 0.89 (0.81 - 0.97) | 11.45% | |
| UiO | 13,315 | 1,105 (8.30%) | 13,315 | 1,422 (10.68%) | 0.71 (0.66 - 0.77) | 29.10% | |
| **META-ANALYSIS** | **112,385** | **6,986 (6.22%)** | **112,385** | **8,745 (7.78%)** | **0.75 (0.64 - 0.86)** | **25.39%** | **0.8739** |
| **COVID-19-related hospitalisation** | | | | | | | |
| SCIFI-PEARL | 32,636 | 67 (0.21%) | 32,636 | 132 (0.40%) | 0.51 (0.38 - 0.68) | 49.41% | |
| SIDIAP | 6,660 | 24 (0.36%) | 6,660 | 55 (0.83%) | 0.43 (0.27 - 0.70) | 56.53% | |
| UiO | 13,445 | 39 (0.29%) | 13,445 | 87 (0.65%) | 0.45 (0.31 - 0.65) | 55.46% | |
| **META-ANALYSIS** | **113,325** | **483 (0.43%)** | **113,325** | **886 (0.78%)** | **0.47 (0.38 - 0.58)** | **52.72%** | **0** |

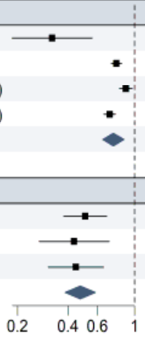

**Supplementary Figure 17:** Vaccine effectiveness estimates for 1st Booster (3rd dose) vs. Primary (2-dose) Vaccination Schema, when follow-up is not stopped at pregnancy end date.

| | Exposed | | Unexposed | | | | |
|---|---|---|---|---|---|---|---|
| Database | Population (N) | Events (N(%)) | Population (N) | Events (N(%)) | HR (95% CI) | VE | i2 |
| **COVID-19 infection** | | | | | | | |
| CPRD GOLD | 1,420 | 98 (6.90%) | 1,420 | 129 (9.08%) | 0.74 (0.57 - 0.96) | 26.12% | |
| SCIFI-PEARL | 21,399 | 873 (4.08%) | 21,399 | 990 (4.63%) | 0.88 (0.80 - 0.96) | 12.38% | |
| SIDIAP | 1,816 | 392 (21.59%) | 1,816 | 447 (24.61%) | 0.83 (0.72 - 0.95) | 17.00% | |
| UiO | 7,706 | 1,339 (17.38%) | 7,706 | 2,355 (30.56%) | 0.48 (0.45 - 0.52) | 51.70% | |
| **META-ANALYSIS** | **64,206** | **5,995 (9.34%)** | **64,206** | **9,324 (14.52%)** | **0.71 (0.50 - 1.02)** | **28.80%** | **0.9765** |
| **COVID-19-related hospitalisation** | | | | | | | |
| SCIFI-PEARL | 22,349 | 43 (0.19%) | 22,349 | 75 (0.34%) | 0.57 (0.39 - 0.83) | 42.74% | |
| SIDIAP | 2,017 | 16 (0.79%) | 2,017 | 13 (0.64%) | 1.23 (0.59 - 2.56) | -22.97% | |
| UiO | 8,782 | 57 (0.65%) | 8,782 | 85 (0.97%) | 0.67 (0.48 - 0.93) | 33.15% | |
| **META-ANALYSIS** | **69,565** | **328 (0.47%)** | **69,565** | **468 (0.67%)** | **0.69 (0.50 - 0.96)** | **30.69%** | **0.3976** |

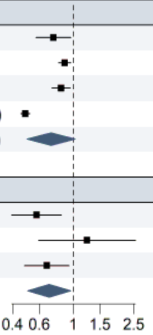

**Supplementary Figure 18:** Hazard Ratios for COVID-19 Infection and COVID-19 related Hospitalisation (Primary (2-dose) Vaccination Schema vs. Unvaccinated, and 1st Booster (3rd dose) vs. Primary (2-dose) Vaccination Schema) stratified by pregnancy trimester at time of vaccination, when follow-up is not stopped at pregnancy end date. Estimates across databases were pooled using random-effects meta-analysis.

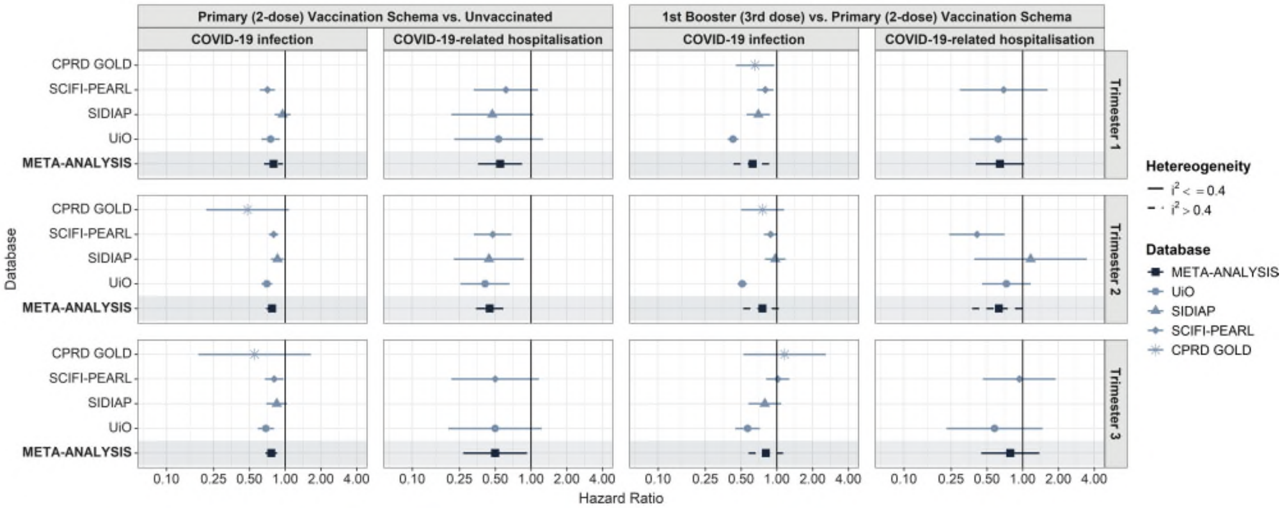

**Supplementary Figure 19:** Hazard Ratios for COVID-19 Infection and COVID-19 related Hospitalisation (Primary (2-dose) Vaccination Schema vs. Unvaccinated, and 1st Booster (3rd dose) vs. Primary (2-dose) Vaccination Schema) stratified by vaccine brand, when follow-up is not stopped at pregnancy end date. Estimates across databases were pooled using random-effects meta-analysis.

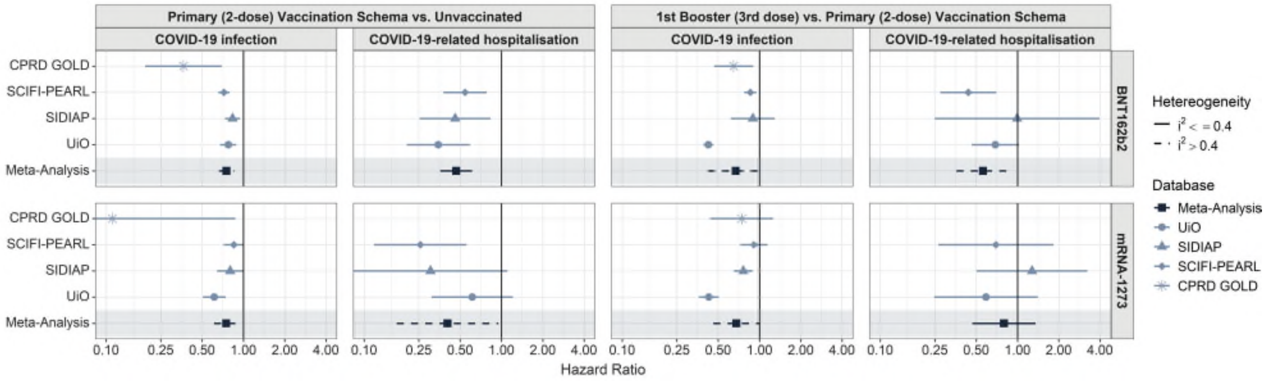

**Supplementary Figure 20:** Hazard Ratios for COVID-19 related Hospitalisation (Primary (2-dose) Vaccination Schema vs. Unvaccinated, and 1st Booster (3rd dose) vs. Primary (2-dose) Vaccination Schema), when hospitalisation related to delivery is considered. Estimates across databases were pooled using random-effects meta-analysis.

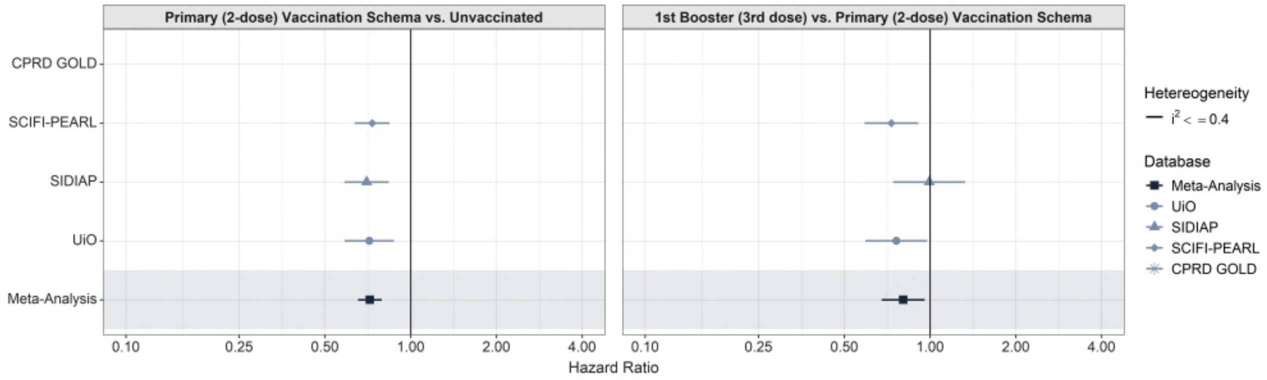

**Supplementary Figure 21:** Vaccine effectiveness estimates for Primary (2-dose) Vaccination Schema vs. Unvaccinated, when COVID-19 records are identified based on polymerase chain reaction (PCR) or antigen postitive tests. Estimates across databases were pooled using random-effects meta-analysis.

| | Exposed | | Unexposed | | | | |
|---|---|---|---|---|---|---|---|
| **Database** | **Population (N)** | **Events (N(%))** | **Population (N)** | **Events (N(%))** | **HR (95% CI)** | **VE** | **i2** |
| **COVID-19 infection** | | | | | | | |
| CPRD GOLD | 2,530 | 9 (0.36%) | 2,530 | 35 (1.38%) | 0.26 (0.12 - 0.53) | 74.34% | |
| SCIFI-PEARL | 32,353 | 925 (2.86%) | 32,353 | 1,203 (3.72%) | 0.76 (0.70 - 0.83) | 24.17% | |
| SIDIAP | 6,502 | 494 (7.60%) | 6,502 | 620 (9.54%) | 0.78 (0.69 - 0.88) | 21.80% | |
| UiO | 13,012 | 455 (3.50%) | 13,012 | 750 (5.76%) | 0.66 (0.59 - 0.74) | 34.23% | |
| **META-ANALYSIS** | **111,456** | **3,595 (3.23%)** | **111,456** | **5,074 (4.55%)** | **0.70 (0.61 - 0.82)** | **29.58%** | **0.7682** |
| **COVID-19-related hospitalisation** | | | | | | | |
| SCIFI-PEARL | 32,550 | 36 (0.11%) | 32,550 | 105 (0.32%) | 0.34 (0.23 - 0.50) | 65.79% | |
| SIDIAP | 6,625 | 18 (0.27%) | 6,625 | 45 (0.68%) | 0.40 (0.23 - 0.69) | 59.86% | |
| UiO | 13,127 | 27 (0.21%) | 13,127 | 80 (0.61%) | 0.38 (0.24 - 0.58) | 62.39% | |
| **META-ANALYSIS** | **112,319** | **195 (0.17%)** | **112,319** | **504 (0.45%)** | **0.37 (0.28 - 0.47)** | **63.45%** | **0** |

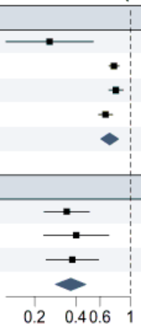

**Supplementary Figure 22:** Vaccine effectiveness estimates for 1st Booster (3rd dose) vs. Primary (2-dose) Vaccination Schema, when COVID-19 records are identified based on polymerase chain reaction (PCR) or antigen postitive tests. Estimates across databases were pooled using random-effects meta-analysis.

| | Exposed | | Unexposed | | | | |
|---|---|---|---|---|---|---|---|
| **Database** | **Population (N)** | **Events (N(%))** | **Population (N)** | **Events (N(%))** | **HR (95% CI)** | **VE** | **i2** |
| **COVID-19 infection** | | | | | | | |
| CPRD GOLD | 1,405 | 69 (4.91%) | 1,405 | 97 (6.90%) | 0.69 (0.51 - 0.94) | 30.63% | |
| SCIFI-PEARL | 21,482 | 726 (3.38%) | 21,482 | 810 (3.77%) | 0.89 (0.80 - 0.98) | 11.08% | |
| SIDIAP | 1,889 | 238 (12.60%) | 1,889 | 284 (15.03%) | 0.81 (0.68 - 0.96) | 18.94% | |
| UiO | 7,778 | 410 (5.27%) | 7,778 | 1,694 (21.78%) | 0.22 (0.20 - 0.24) | 78.25% | |
| **META-ANALYSIS** | **64,769** | **2,862 (4.42%)** | **64,769** | **6,882 (10.63%)** | **0.57 (0.25 - 1.29)** | **42.69%** | **0.9923** |
| **COVID-19-related hospitalisation** | | | | | | | |
| SCIFI-PEARL | 22,406 | 21 (0.09%) | 22,406 | 56 (0.25%) | 0.37 (0.23 - 0.62) | 62.64% | |
| SIDIAP | 2,041 | 10 (0.49%) | 2,041 | 12 (0.59%) | 0.84 (0.36 - 1.94) | 16.13% | |
| UiO | 8,664 | 25 (0.29%) | 8,664 | 49 (0.57%) | 0.52 (0.32 - 0.85) | 47.75% | |
| **META-ANALYSIS** | **69,453** | **123 (0.18%)** | **69,453** | **245 (0.35%)** | **0.50 (0.34 - 0.74)** | **50.18%** | **0.2773** |

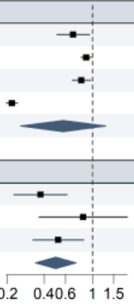
